# Study of effect modifiers of genetically predicted CETP reduction

**DOI:** 10.1101/2021.09.09.21263362

**Authors:** Marc-André Legault, Amina Barhdadi, Isabel Gamache, Audrey Lemaçon, Louis-Philippe Lemieux Perreault, Jean-Christophe Grenier, Marie-Pierre Sylvestre, Julie G. Hussin, David Rhainds, Jean-Claude Tardif, Marie-Pierre Dubé

## Abstract

Genetic variants in drug targets can be used to predict the effect of drugs. Here, we extend this principle to assess how sex and body mass index may modify the effect of a genetically predicted lower CETP levels on biomarkers and cardiovascular outcomes.

We found sex and BMI to be modifiers of the association between genetically predicted lower CETP and lipid biomarkers in UK Biobank participants. Female sex and lower BMI were associated with higher HDL-cholesterol and lower LDL-cholesterol for a same genetically predicted reduction in CETP concentration. We found that sex also modulated the effect of genetically lower CETP on cholesterol efflux capacity in samples from the Montreal Heart Institute Biobank. However, these modifying effects did not extend to sex-differences in cardiovascular outcomes in our data.

Our results provide insight on the clinical effects of CETP inhibitors in the presence of effect modification based on observational genetic data. The approach can support precision medicine applications and help assess the external validity of clinical trials.

## Introduction

Genetic variants in drug targets can be used to predict the effects of drugs ^1,2^. The identification of rare variants with strong effects on protein function led to the development of new drug classes, and there is a growing number of genetically supported drug targets along various phases of drug development ^3–5^.

However, few genetic studies of drug targets have focused on the identification of subgroups of individuals that could derive a greater benefit from the drug. This question is central to our quest to improve precision medicine, which can be supported by genetic techniques. Randomized controlled trials (RCTs) are powered to detect the benefit of an intervention in the full study population and analyses in subgroups of individuals are typically reported as exploratory observations. When clinical or demographic subgroups are underrepresented or excluded from trials, external validity can be put into question ^6^. In clinical trials of cardiovascular disease prevention, for example, women are frequently underrepresented, and only 31% of trials report sex-specific results ^7^. This may be of importance as differences in body size and composition as well as hormonal and gender differences could all have an impact on drug response ^8^.

CETP inhibitors have a complex history of heterogeneous findings from RCTs and genetic studies. Three of the four trials of CETP inhibitors did not report benefit, except for the most recent and largest trial (Randomized EValuation of the Effects of Anacetrapib through Lipid-modification, REVEAL) that showed a small reduction in risk of cardiovascular outcomes in an at-risk population, with a rate ratio of 0.91 (95% CI 0.85, 0.97) for the study primary efficacy endpoint ^9^.

In this paper we investigate how sex and body mass index (BMI) may modify the effect of a genetically predicted CETP reduction on biomarkers and cardiovascular outcomes. We consider the effect on apolipoproteins and lipid fractions thought to be related to CETP inhibition, namely high-density lipoprotein cholesterol (HDL-c), low-density lipoprotein cholesterol (LDL-c), and their main lipoprotein constituents: apolipoproteinA (apoA) and apolipoproteinB (apoB), respectively. We also consider C-reactive protein levels, a measure of systemic inflammation, and lipoprotein(a) an independent atherogenic lipoprotein which may be affected by CETP inhibition. We also assessed the effect of genetically predicted CETP reduction on the capacity of apoB-depleted plasma to efflux cholesterol using an independent dataset from the Montreal Heart Institute (MHI) Biobank. This work is important both for its contribution to precision medicine methodological applications, and to gain a better understanding of the role of CETP in lipid homeostasis and disease pathology. Our approach is akin to a Mendelian Randomization study, and a justification for the causal interpretation is presented in the Supplementary Methods.

## Methods

### Study populations

Data from the UK Biobank cohort were used for the analyses relating genetically predicted CETP with biomarkers and cardiovascular outcomes. The variable definitions and genetic quality control steps are described in the Supplementary Methods.

Data from the MHI Biobank cohort were used to conduct the analyses of genetically predicted CETP with cholesterol efflux (Supplementary Methods).

### Genetic predictors of CETP activity

We evaluated various genetic scores of CETP activity based on a GWAS of plasma CETP concentration or the MAGNETIC nuclear magnetic resonance GWAS ^10,11^ constructed using the p-value thresholding and LD clumping method (Supplementary Methods). All the genetic scores were highly correlated with r^2^ between 0.75 and 1.00 (Supplementary Table 1), and for subsequent analyses, we selected the score based on the plasma CETP concentration as it directly relates to CETP activity.

Given that genetic scores and variants have complementary advantages, we also conducted analyses using the rs1800775 variant. The *CETP* variant -629C>A (rs1800775) is known to disrupt transcription factor binding and to reduce CETP activity ^12,13^. Using such a variant as a proxy for CETP levels has the advantage of providing interpretable results in allelic units and does not rely on weights which may be biased with respect to the population (both in terms of sex, clinical profile and ethnicity) in which they were estimated. For example, if genetic variants have different effects in men and women, scores based on these effects will be better predictors of the phenotype in individuals whose sex was most prevalent in the study from which the estimates are derived.

Because this study investigates the effect of genetically predicted CETP levels, it is important to validate the statistical strength of our predictors. We estimated association strength using HDL-c and LDL-c levels as proxy variables because they are known to be impacted by CETP modulation. We used univariable linear regression and we report F statistics and R^2^ as is usual in Mendelian randomization studies ^14^. When regressing on measured HDL-c in the UK Biobank, the F statistic was 10,400 (F_1;362,466_) for the genetic score and 7,239 (F_1;362,466_) for rs1800775, with corresponding R^2^ of 0.028 and 0.020, respectively. Similarly, when regressing on LDL-c, the F statistics and R^2^ were 201 (F_1;394,285_) and R^2^ =0.00051 for the genetic score and 160 (F_1;394,285_) and R^2^=0.00041 for rs1800775. The genetic score has a stronger effect than rs1800775 on CETP and consequently on HDL-c and LDL-c. The rs1800775 variant remains a strong predictor of CETP and is not prone to bias due to weighting.

### Statistical Analyses

We assessed the effect of the genetic predictors of CETP on observed biomarkers and cardiovascular outcomes using linear and logistic regression models as appropriate. To estimate the effect modification by sex and BMI, we fit the models containing a product interaction term between the genetic predictor of CETP and the effect modifier of interest and fixed covariates including the component variables of the interaction and age, sex and ancestry principal components. We adjusted for these covariates to improve the precision of our estimates. We used hypothesis testing of a null product interaction coefficient as a test of effect modification and the p-value for this test is denoted as *p*_*itx*_. In linear models, the hypothesis test is for an additive interaction and it can be interpreted as an additive deviation from the contribution of the interacting variables. In logistic regression models, the test of the product interaction term assesses a multiplicative effect modification of the odds ratio (Supplementary Methods). In other words, in the latter case the test is of OR_11_ / (OR_01_ × OR_10_) = 1 where OR_ij_ denotes the odds ratio when setting the first interacting variable to *i* and the 2^nd^ variable to *j* compared to the reference value for both covariables (Supplementary Methods). Using the logistic regression model, we also computed interaction statistics on the additive scale namely the Relative Excess Risk due to Interaction (RERI) and the interaction contrast on the probability scale (Supplementary Methods). The additive effect statistics were developed for risk factors (and not preventive exposures) which prompted us to report interaction effects for the “male” sex, for a 1 s.d. increase in BMI and for a 1 s.d. increase in the CETP score. To facilitate interpretation, we also used interaction models to compute the marginal effects of the genetic CETP predictors on the outcomes of interest at representative values of the tested modifiers. In models with more than a single interaction term, we used the R package “margins” to estimate the marginal effect at representative cases and corresponding 95% confidence interval, otherwise we computed the marginal effects directly by summing the relevant regression coefficients.

In order to test for possible nonlinear effects of BMI and the CETP genetic score (and their interaction), we fitted an interaction model with interacting restricted cubic splines with four knots for BMI and the CETP genetic score. We used ANOVA to test for interactions by simultaneously considering all interaction coefficients. For significant interactions in the nonlinear models, we then plotted the predicted effects at varying levels of BMI and of the CETP genetic score to visualize the nonlinear effects. We used the R ‘rms’ package to conduct these analyses.

For analyses based on the CETP genetic score, all effects for biomarkers are reported in units of standard deviation (s.d.) of the biomarker per s.d. decrease of the score (representing a decrease in CETP concentration as for pharmacological CETP inhibition). For binary outcomes, the reported effects are odds ratios per s.d. decrease of the genetic score. For analyses based on the individual CETP variant, the results are expressed per copy of the “A” allele at rs1800775 (CETP -629C>A) which decreases CETP levels. For causal interpretation of these analyses, see Supplementary Methods. No adjustment was made for multiple testing of phenotypes and effect modifiers. Estimates are reported with 95% confidence intervals.

### Power Analyses

We estimated the power of the different association models using simulations. In the simplest case, we simulated a normally distributed genetic score with a fixed effect on a standard normal outcome and computed the proportion of rejected null hypotheses across simulation replicates at *α* = 0.05. We extended this model to account for interaction effects, and we used a latent variable logistic regression model when estimating the power for association with binary traits. The simulation model is described in Supplementary Methods.

## Results

### Study population

There were 413,138 unrelated participants of European origin from the UK Biobank included in the analyses (Supplementary Methods). The number of events under consideration for the cardiovascular outcomes as well as the descriptive statistics for continuous measurements are presented in Table 1.

**Table 1.** Descriptive statistics of the effect modifiers, biomarkers and cardiovascular outcomes in the UK Biobank study population.

|  | Women | Men | All |
| --- | --- | --- | --- |
| **General characteristics** |  |  |  |
| n (%) | 222,684 (54%) | 190,454 (46%) | 413,138 |
| age - mean ± s.d. | 56.6 ± 7.88 | 57.0 ± 8.06 | 56.8 ± 7.96 |
| **Effect modifiers** |  |  |  |
| Sex - n (%) | 222,684 (100%) | 190,454 (100%) | 413,138 (100%) |
| Body mass index (original scale) (BMI, kg/m<sup>2</sup>) - Mean ± s.d. (median ± IQR) | 27.0 ± 5.15<br>(26.1 ± 6.21) | 27.9 ± 4.24<br>(27.3 ± 5.07) | 27.4 ± 4.77<br>(26.7 ± 5.74) |
| Body mass index (BMI, ln(kg/m<sup>2</sup>)) - Mean ± s.d. | 3.28 ± 0.18 | 3.32 ± 0.15 | 3.30 ± 0.17 |
| **Biomarkers as outcomes** |  |  |  |
| Lipoprotein(a) (lp(a), nmol/L) - Mean ± s.d. | 44.6 ± 49.4 | 43.4 ± 49.3 | 44.0 ± 49.4 |
| Apolipoprotein B (apoB, g/L) - Mean ± s.d. | 1.04 ± 0.24 | 1.03 ± 0.24 | 1.03 ± 0.24 |
| Low-density lipoprotein cholesterol (LDL-c, mmol/L) - Mean ± s.d. | 3.64 ± 0.87 | 3.48 ± 0.86 | 3.57 ± 0.87 |
| Apolipoprotein A (apoA, g/L) - Mean ± s.d. | 1.64 ± 0.27 | 1.43 ± 0.23 | 1.54 ± 0.27 |
| High-density lipoprotein cholesterol (HDL-c, mmol/L) - Mean ± s.d. | 1.60 ± 0.38 | 1.29 ± 0.31 | 1.45 ± 0.38 |
| C-reactive protein (original scale) (CRP, mg/L) - Mean ± s.d.<br>(median ± IQR) | 1.43 ± 2.97<br>(1.37 ± 2.30) | 1.36 ± 2.77<br>(1.29 ± 1.88) | 1.39 ± 2.89<br>(1.33 ± 2.09) |
| C-reactive protein - (CRP, ln(mg/L)) - Mean ± s.d. | 0.36 ± 1.09 | 0.30 ± 1.02 | 0.33 ± 1.06 |
| **Cardiovascular outcomes** |  |  |  |
| Myocardial infarction (MI) - n (%) | 4,747 (2%) | 13,812 (7%) | 18,559 (4%) |
| Percutaneous coronary intervention or coronary artery bypass graft (PCI/ CABG) - n (%) | 3,395 (2%) | 13,546 (7%) | 16,941 (4%) |
| Coronary artery disease, soft (CAD) - n (%) | 14,803 (7%) | 29,910 (16%) | 44,713 (11%) |
| Coronary artery disease, hard (CAD) - n (%) | 6,825 (3%) | 19,517 (11%) | 26,342 (7%) |

*If a transformation was needed to normalize the data, the statistics are given in both the original and transformed scale. The units represent the applied transformation.*

### Effect of genetically predicted reduction of CETP on biomarkers and cardiovascular outcomes

We first assessed the effect of the genetic CETP predictors on biomarkers and cardiovascular outcomes without any modifiers to contextualize future results (Table 2). As expected, the strongest association was with HDL-c, with an increase of 0.167 s.d. in HDL-c (corresponding to 0.064 mmol/l) per 1 s.d. decrease of the CETP genetic score, with concordant results for the rs1800775 (CETP -629C>A) SNP alone (Supplementary Table 3). To offer a comparison, pharmacological CETP inhibition with anacetrapib increased HDL-c levels by 1.11 mmol/l on average at trial midpoint, evacetrapib increased HDL-c levels by 1.52 mmol/l at 3 months whereas dalcetrapib increased HDL-c by about 0.34 mmol/l on average at 1 year ^9,15,16^. In these examples, the pharmacological effect of CETP inhibition is about 17x stronger for anacetrapib, 24x for evacetrapib and 5x for dalcetrapib when compared to a 1 s.d. reduction of the CETP genetic score. This comparison is based only on the effect on HDL-c levels which may not represent the full spectrum of effects of CETP inhibitors.

**Table 2.** Association of the CETP genetic score with biomarkers and cardiovascular events.

| Biomarkers - in s.d. units | N | Standardized coefficient * | p-value |
| --- | --- | --- | --- |
| Lipoprotein(a) | 315,214 | -0.011 (-0.013, -0.008) | $1.0 \times 10^{-14}$ |
| C-reactive protein (CRP) | 394,165 | 0.0026 (-0.0005, 0.0057) | 0.098 |
| HDL cholesterol | 362,468 | 0.167 (0.164, 0.170) | $<10^{-300}$ |
| Apolipoprotein A | 360,451 | 0.131 (0.129, 0.134) | $<10^{-300}$ |
| LDL cholesterol | 394,287 | -0.023 (-0.026, -0.019) | $1.4 \times 10^{-45}$ |
| Apolipoprotein B | 393,089 | -0.033 (-0.036, -0.030) | $5.1 \times 10^{-97}$ |
| Basal cholesterol efflux (MHI Biobank) | 5,215 | 0.105 (0.079, 0.130) | $1.9 \times 10^{-15}$ |
| cAMP-stimulated cholesterol efflux (MHI Biobank) | 5,214 | 0.085 (0.059, 0.111) | $2.7 \times 10^{-10}$ |
| Cardiovascular outcomes | N cases / N total | Odds ratio (95% CI) * | p-value |
| Coronary artery disease (“soft”) | 44,713 / 413,138 | 0.975 (0.965, 0.985) | $8.4 \times 10^{-7}$ |
| Coronary artery disease (“hard”) | 26,342 / 394,767 | 0.971 (0.958, 0.983) | $6.0 \times 10^{-6}$ |
| Myocardial infarction | 18,559 / 413,138 | 0.980 (0.966, 0.995) | 0.0096 |
| Percutaneous coronary intervention or coronary artery bypass graft | 16,941 / 413,138 | 0.967 (0.952, 0.982) | $2.6 \times 10^{-5}$ |

\* Coefficients for continuous variables (biomarkers) are from a linear regression model adjusted for age, sex and the first 10 principal components and are expressed in standard deviation units of the biomarkers outcome per standard deviation reduction in the CETP genetic score. Odds ratios for cardiovascular outcomes are estimated using a logistic regression model adjusted for the same covariates.

The CETP genetic score was strongly associated with both basal and cAMP-stimulated cholesterol efflux capacity as measured in plasma from 5,215 participants of the MHI Biobank (Table 2). A 1 s.d. decrease in the score increased basal cholesterol efflux by 0.105 s.d. (95% CI 0.078, 0.130) and increased cAMP-stimulated cholesterol efflux by 0.085 s.d. (95% CI 0.059, 0.011).

The CETP genetic score was associated with LDL-c levels with a 0.023 s.d. decrease in LDL-c (corresponding to 0.020 mmol/l) per 1 s.d. decrease of the CETP genetic score (p = 1.4 × 10^−45^). Most RCTs of CETP inhibitors reported a decrease in LDL-c cholesterol, but not in the dal-OUTCOMES trial of dalcetrapib. Recently, anacetrapib was shown to decrease the production of lp(a) which could partly explain the benefit of this CETP inhibitor ^17,18^. A decrease in lp(a) levels was also observed with torcetrapib suggesting a possible class effect of CETP inhibitors ^19^. We tested the association between the CETP genetic score and lp(a) levels measured in UK Biobank participants. A reduction of 1 s.d. in the CETP genetic score was associated with a decrease in lp(a) levels by 0.011 s.d. (95% CI 0.008, 0.013);p = 1.0 × 10^−14^. In laboratory units of lp(a) levels, this corresponds to 0.524 nmol/l of lp(a) per s.d. of the CETP genetic score.

The association of the CETP genetic score with cardiovascular outcomes was concordant with the observed associations with the lipid profile. One s.d. reduction in the CETP genetic score was associated with CAD (“soft” definition, Supplementary Table 2) with an OR of 0.97 (95% CI 0.96, 0.98) p = 8.4 × 10^−7^. Previous studies of genetic CETP reduction have also reported effects scaled by a 10 mg/dL reduction in apoB levels with an OR of 0.78 (95% CI 0.71, 0.86) ^20^. After scaling for an effect of this magnitude, our estimate corresponded to an OR of 0.72 (95% CI 0.64, 0.82). We also repeated these analyses for the rs1800775 *CETP* promoter variant and obtained similar results (Supplementary Table 3).

### Female sex is associated with larger benefit of genetically lower CETP on the lipid profile

The large phase III RCTs of CETP inhibitors suffered from large sex imbalances, ranging between 16% of female participants (REVEAL) and 23% (ACCELERATE) and it is unlikely that these trials could have identified effect differences between men and women. In Figure 1, we show the reported drug effects from the major RCTs of CETP inhibitors stratified by sex. We conducted an inverse variance-weighted meta-analysis of the effect of the CETP inhibitor in the dal-OUTCOMES, REVEAL and ACCELERATE studies (Supplementary Methods). We did not include the ILLUMINATE trial with torcetrapib, as the drug had off-target deleterious effects. The calculated meta-analysis risk ratio is 0.96 (95% CI 0.91, 1.02) in men and 0.92 (95% CI 0.82, 1.04) in women. The test for heterogeneity between the male and female effects was not significant (p = 0.50). The Cochran Q statistics for heterogeneity between studies were 5.5 (p = 0.063) for the overall effect, 0.025 (p = 0.99) for the female-specific effect and 8.0 (p = 0.018) for the male-specific effect.

**Figure 1.**
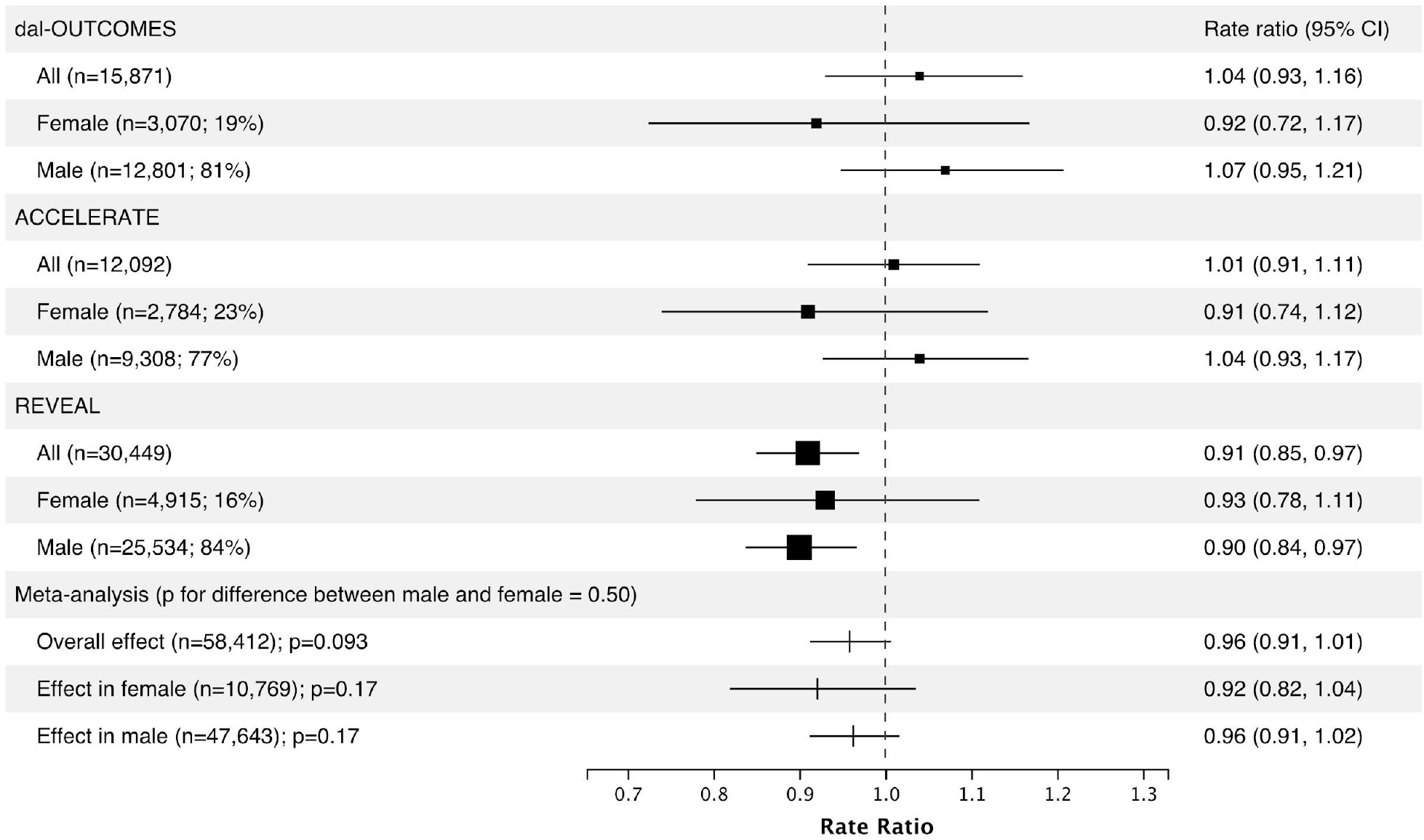
Effect of treatment from phase 3 trials of CETP inhibitors in the whole population and stratified by sex. Points are scaled with respect to the relative weight in the overall and sex-specific meta-analyses.

We used regression models to assess the interaction effect of sex with the CETP genetic score on biomarkers and cardiovascular outcomes in the UK Biobank (Figure 2). We observed statistically significant interactions for apoA and apoB as well as LDL-c and HDL-c levels. The strongest effect modification was with HDL-c and apoA. For instance, a one s.d. unit reduction in the CETP score increased HDL-c by 0.15 s.d. (95%CI 0.14, 0.15) in men (p < 10^−300^) and by 0.18 s.d. (95% CI 0.18, 0.19) in women (p < 10^−300^) and the interaction p-value was 5 × 10^−32^. In general, genetically predicted lower CETP had a more beneficial effect on the lipid profile in women than men. Similar results were obtained with the rs1800775 variant alone (Supplementary Figure 1).

**Figure 2.**
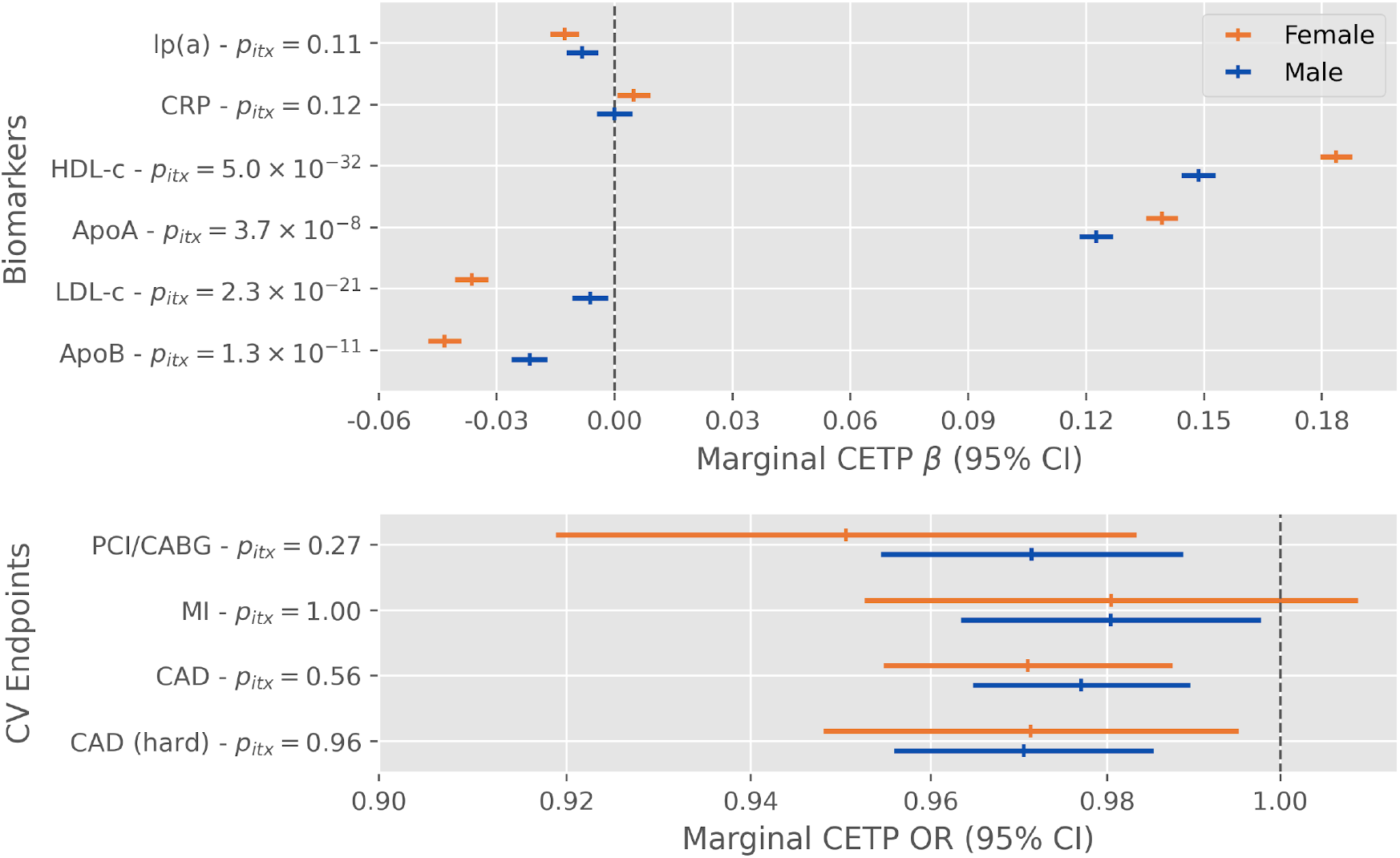
Effect modification of a 1 standard deviation decrease in the CETP concentration genetic score by sex on biomarkers and cardiovascular outcomes in the UK Biobank. Displayed p-values (p_itx_) are for the two-sided test of the product interaction term between the CETP score and a binary sex indicator variable.

We tested for the interaction of sex with the CETP genetic score on cholesterol efflux measured in a subgroup of participants of the MHI Biobank. For cAMP-stimulated cholesterol efflux, a unit decrease in the score was associated with a 0.064 s.d. (95% CI 0.032, 0.095) increase in efflux for men (p = 7 × 10^−5^) and 0.13 (95% CI 0.086, 0.18) for women (p = 5 × 10^−8^) and the interaction p-value was 0.02. A similar effect was also observed for basal efflux (Supplementary Figure 2). Again, women had a more favourable cholesterol efflux profile than men, with lower genetically-predicted CETP levels, as their cholesterol efflux capacity was higher on average.

We considered whether the differences of the effect of CETP on biomarkers according to sex also led to differences in cardiovascular outcomes. On the multiplicative scale, the difference in the effect of a genetic CETP reduction between men and women was not statistically significant (interaction p-value of 0.56 for CAD). However, the RERI (0.082 95% CI [0.020, 0.146]) and interaction contrast (0.0017 95% CI [0.00024, 0.0032]) suggested a difference in the effect of the CETP score on CAD (“hard”) in men compared to women (Supplementary Table 5). Power analyses rule out a strong effect difference of genetically lower CETP concentration on cardiovascular outcomes between men and women (Supplementary Appendix, Supplementary Figure 3). In sensitivity analyses, we show that results are robust to further adjustment for statin use (Supplementary Figure 4).

### Higher BMI reduces the benefit of genetically lower CETP on the lipid profile

Previous studies have reported that the effect of CETP on HDL-c is different across BMI class ^21,22^. To assess how the effect of genetic CETP modulation is affected by BMI, we used interaction models from which we draw inferences and report marginal effects at fixed BMI values (Figure 3). We found BMI to be a significant modulator of the association between the CETP genetic score and HDL-c (interaction p = 5.4 × 10^−73^) and apoA levels (interaction p = 5.2 × 10^−24^) in the UK Biobank. Individuals with lower BMI had higher HDL-c and apoA levels per 1 s.d. decrease in the genetic CETP score. BMI was also a modulator of the association between the CETP genetic score and LDL-c (interaction p = 0.00013) and apoB levels (interaction p = 0.0012) with a lower BMI being associated with lower levels of LDL-c and apoB per s.d. decrease of the CETP genetic score (Figure 3). The interaction term was also significant for lp(a) supporting that BMI is also a modulator of the association between the CETP genetic score and lp(a) (p = 0.0027, Figure 3). In individuals of normal BMI, one s.d. reduction in the genetic CETP score was associated with 0.018 s.d. lower lp(a) (95% CI 0.014, 0.023) with p = 8 × 10^−15^ and individuals of obese BMI had 0.0081 s.d. lower lp(a) (95% CI 0.0025, 0.014) with p = 0.004 (Supplementary Figure 5). The interaction between the genetic CETP score and BMI was not significant for C-reactive protein (p = 0.24). In the MHI Biobank, there was no interaction between BMI and the genetic CETP score on basal and cAMP-stimulated cholesterol efflux (p = 0.31 and p = 0.84, respectively). Results based on the rs1800775 variant were concordant with those based on the genetic CETP score (Supplementary Figure 6).

**Figure 3.**
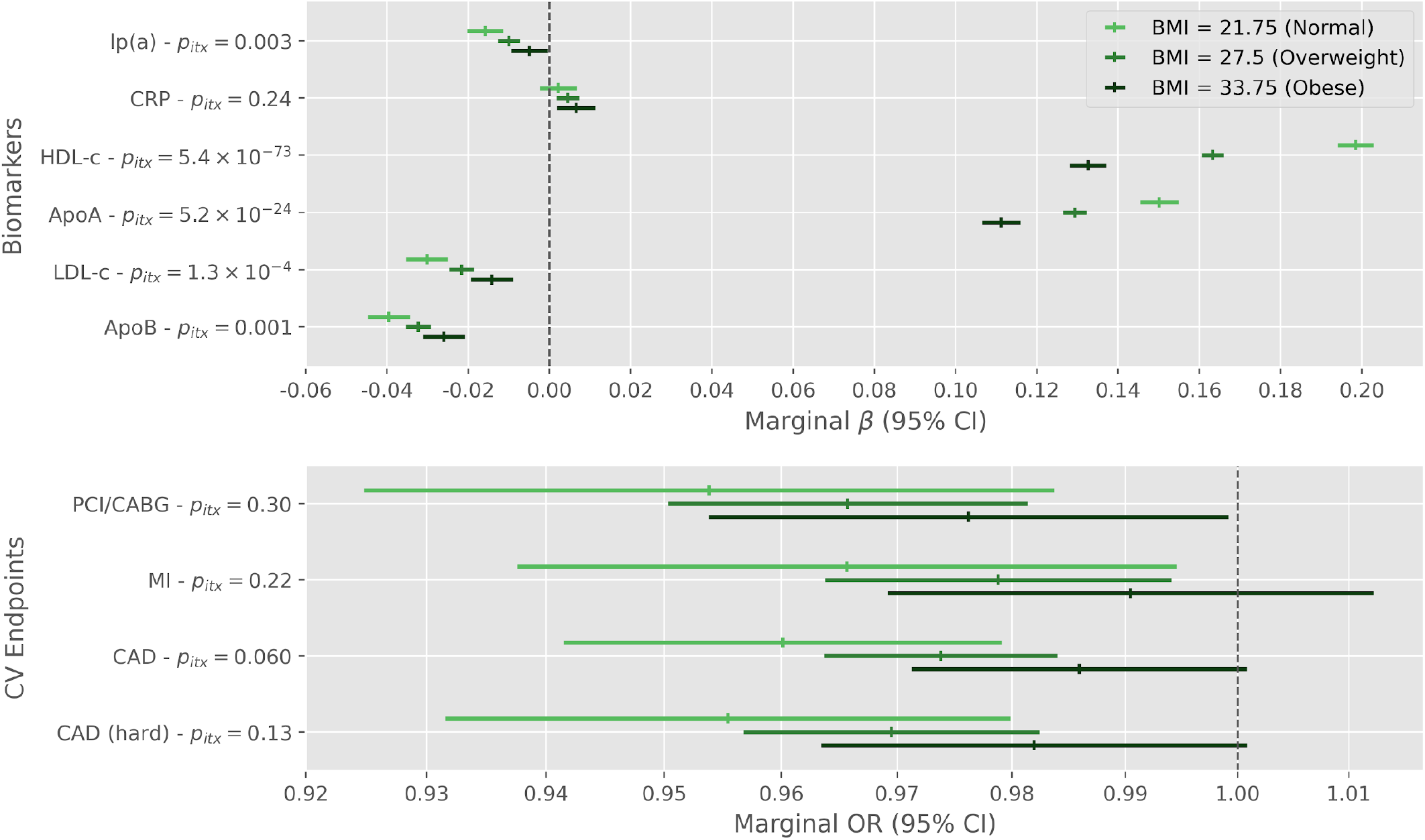
Effect modification by body mass index of a 1 standard deviation decrease in the CETP concentration genetic score on biomarkers and cardiovascular outcomes in the UK Biobank. Displayed p-values (p_itx_) are for the two-sided test of the product term between the CETP score and standardized body mass index.

To allow for non-linear effects of BMI and the CETP genetic score, we used linear and logistic regression with interacting restricted cubic splines to model these two variables. There was evidence of nonlinear effects for BMI on all considered cardiovascular outcomes and biomarkers except for the cholesterol efflux (Supplementary Table 6). The CETP genetic score exhibited possible nonlinear effects on LDL-c (p = 0.0013) and HDL-c (p < 0.0001) and their associated lipoproteins. When considering all linear and nonlinear interaction terms (9 degrees of freedom test), there was evidence for interaction between the CETP score and BMI for lipoprotein(a) levels (p = 0.0233), HDL-c (p < 0.0001), apolipoprotein A (p < 0.0001), LDL-c (p = 0.0001), and apolipoprotein B (p = 0.0147). There was no statistically significant nonlinear interaction with any of the cardiovascular outcomes. To facilitate the interpretation of the nonlinear interactions for biomarkers, we plotted the predicted value of the standardized outcome while varying the levels of the CETP genetic score and BMI (Supplementary Figure 7).

We tested the modulatory effect of BMI on the relationship between the CETP genetic score and cardiovascular outcomes. None of the tested outcomes had statistically significant interactions between BMI and the CETP genetic score, but the effects were directionally consistent with the effects on the lipid profile in the multiplicative scale (Figure 3, Supplementary Table 7). For instance, the interaction coefficient p-value for CAD was 0.060 and the marginal OR of the CETP score on CAD was 0.960 (95% CI 0.941, 0.980) when fixing BMI at 21.75 (normal) versus 0.986 (95% CI 0.971, 1.00) when fixing BMI at 33.75 (obese). We determined the minimum detectable interaction using power analyses (Supplementary Appendix, Supplementary Figure 8). Similar results are observed in subgroups of individuals based on their BMI class (Supplementary Figure 5).

In sensitivity analyses, we show that the modulatory effects of BMI are robust to further adjustment for type II diabetes, ruling out the possibility of mediation of the BMI modulatory effect through diabetes (Supplementary Figure 9).

Because both BMI and sex were important modifiers of the effect of CETP on biomarkers, we evaluated the possibility of a three-way interaction between sex, BMI and the CETP genetic score. There were sex differences in the interaction between BMI and genetic CETP levels on cholesterol efflux (three-way interaction p_itx_ = 0.0037 and p_itx_ = 0.007 for basal and cAMP-stimulated efflux, respectively), LDL-c (p_itx_ = 0.00041) and apoB levels (p = 0.001) (Supplementary Figure 10-13). These results are further described in the Supplementary Appendix.

## Discussion

Using the large UK Biobank resource, supported by cholesterol efflux measurements in the MHI Biobank, we report the effect of genetically lower CETP on lipid biomarkers, cholesterol efflux, CRP, and cardiovascular outcomes. We assessed how sex and BMI changed the effect of a genetically predicted decrease of CETP on those measurements and outcomes. We report significant modulatory effect of both sex and BMI on the association of CETP with biomarkers, but we were unable to show that these differences resulted in an effect on cardiovascular outcomes.

In our analyses, we observed that a genetically predicted lower CETP concentration was strongly associated with higher HDL-c and apoA levels and, to a lesser extent, to lower LDL-c and apoB levels. These results are concordant with previous reports of the effect of CETP on lipids and lipoproteins ^23,24^. We also observed lp(a) levels to be slightly, but significantly, lower in individuals with a genetically predicted reduction in CETP. This observation is interesting as lp(a) is an important risk factor for CAD that is largely independent of other lipoproteins. Previous studies have reported that both torcetrapib and anacetrapib could reduce lp(a) levels ^17,19,25^. The added genetic support could be indicative of a class effect of CETP inhibitors. In a Mendelian randomization study of lp(a) levels, a 10 mg/dl genetic reduction in lp(a) was associated with an OR of 0.942 for coronary heart disease supporting the causal role of lp(a) in coronary heart disease ^26^. In our study, we show that a 1 s.d. decrease in the genetic CETP score was associated with a reduction in lp(a) of about 2 mg/dl which corresponds to an OR for coronary artery disease of 0.988 based on this previous MR study.

Genetically lower CETP was not associated with C-reactive protein. In the dal-OUTCOMES trial of dalcetrapib and the ACCELERATE trial of evacetrapib, CETP inhibition was associated with an increase in C-reactive protein, but there was no significant difference in the DEFINE trial of anacetrapib ^15,16,27^. Given our high power to detect an association with biomarker measurements in the UK Biobank, it is unlikely that a lifelong, genetically lower CETP level has an effect on C-reactive protein levels.

We have also found that genetically lower CETP was associated with higher levels of cholesterol efflux. This observation is concordant with previous reports of increased cholesterol efflux in patients treated with dalcetrapib and anacetrapib ^28,29^. Genetically lower CETP was also associated with lower rates of cardiovascular outcomes, with 2.5% fewer CAD events per s.d. decrease in the CETP genetic score. The observed protective effect was robust to adjustment for observed apoB levels at baseline in the UK Biobank, supporting that the protective effect of CETP may not be exclusively mediated by apoB levels. However, adjusting for observed apoB levels at a single time point may not completely control for a lifetime reduction in apoB levels by CETP genetic variants. Nonetheless, we presented a unified portrait of the effect of genetically lower CETP which may help better understand on-target effects of CETP inhibitors and the diversity of pathways through which genetic variants in *CETP* may exert a protective effect on CAD.

The value of using human genetic variants to predict the effect of pharmacological modulation of drug targets is gaining recognition as more examples of drug target discoveries and predictions for ongoing randomized trials are reported ^2,30^. Human genetics can also inform on subgroup effects in the context of precision medicine, for clinical trial design, or to estimate external validity of drug effects to other patient populations. Genetic studies of *CETP* variants have highlighted possible effect modification by sex on HDL-c levels, carotid intima-media thickness, the HDL-c / apoAI ratio and on the dynamics of postprandial triglyceride levels ^31–35^. Sex differences were also observed in many traits thought to be involved in the atheroprotective effect of CETP such as the apoAI and apoA-II composition of HDL and CETP mediated cholesterol efflux ^36,37^. Whether this translates to cardiovascular outcomes remains unknown ^38^. Considering that female sex is underrepresented in the majority of cardiovascular clinical trials, sex differences may have important clinical implications as inferences drawn from the unbalanced trial populations are used to inform treatment.

In our analyses, we found sex to be a strong modulator of the effect of genetically lower CETP on lipid biomarkers and cholesterol efflux. Women with CETP lowering genetic variants had lower LDL-c and apoB levels and higher HDL-c, apoA and cholesterol efflux. In a substudy of the DEFINE trial, anacetrapib increased cholesterol efflux in men, but not women ^29^. In that study, cholesterol efflux capacity was measured using fluorescently labeled cholesterol which mostly captures efflux through the ABCA1 pathway. Here, we used radiolabeled cholesterol and our measurements in cAMP-stimulated conditions also include the contributions of ABCG1 and SR-BI. In healthy subjects, serum from women had more SR-BI mediated efflux capacity whereas serum from men had more ABCA1-mediated efflux capacity ^39^. We suggest that, on average, with genetically lower CETP, women have more overall cholesterol efflux capacity than men, but that specific pathways may show the inverse relationship. The modulatory effect of sex did not translate to strong differences in cardiovascular outcomes in our analysis even though the additive interaction model suggests that men had a larger relative risk of “hard” CAD than women for a same increase in the CETP score. Because the UK Biobank contains relatively few cardiovascular events, especially in women, and the genetic variants in *CETP* have a limited effect size, a replication from a large, well-powered study is warranted to confirm these findings. A previous study also reported sex differences in the association between CETP genetic variants and cardiovascular disease^40^, but sex interaction was not formally assessed and the sample size and power may have been limited in that study (n=866).

We conducted a meta-analysis of the sex-stratified results of three RCTs of CETP inhibitors and although we did observe a nominally greater protective effect in women (RR=0.92) than in men (RR=0.96), the difference in effect was not statistically significant (p = 0.50). A total of 10,769 women pooled across all three studies were included in the meta-analysis for a total of 58,412 participants, which is still lower than the number of individuals included in the smallest study (n = 12,092 for ACCELERATE), suggesting limited power to identify a subgroup effect in women. We conclude that there is some indication of a stronger beneficial effect of CETP inhibition in women, but that confirmation using genetic datasets enriched for cardiovascular events or clinical trials with a greater representation of women would be needed.

Plasma CETP is mainly secreted by macrophages in the liver (Kupffer cells) and adipose tissue does not appear to be a clinically relevant contributor to circulating CETP levels ^41,42^. However, higher BMI may affect hepatic and systemic inflammation and result in changes in lipid homeostasis that could alter the function of CETP without affecting plasma CETP concentration. There is also previous evidence of an interaction between obesity status and a CETP variant on acute coronary syndrome from a smaller study (n=474) ^43^.

In our analyses, the atheroprotective profile of lipoproteins attributable to genetically lower CETP levels was stronger in individuals with lower BMI, when compared to individuals with higher BMI, showing lower levels of LDL-c, apoB and lp(a) and higher levels of HDL-c and apoA with genetically lower CETP. Similar results were observed in models allowing for nonlinear effects on the biomarkers. The modulatory effect of BMI on the relationship between CETP and biomarkers did not translate to cardiovascular outcomes, however, a larger dataset may be needed to assess the possible impact on outcomes with sufficient power.

We also found evidence of three-way interactions of sex, BMI and genetically predicted CETP on LDL-c, apoB levels and cholesterol efflux. The attenuated effect of lower CETP on LDL-c reduction with increasing BMI was specific to men. In women, the increase in cholesterol efflux by genetic reduction in CETP was attenuated with increasing BMI, but this effect was sex-specific.

Our study had some limitations. We relied on common genetic variants to model pharmacological CETP inhibition, but these variants do not include rare mutations which can have much stronger effects on CETP function. CETP activity can also be modulated in more subtle ways than complete inhibition, with molecules such as dalcetrapib preserving pre-β-HDL formation, a function that is inhibited by anacetrapib ^44^. This effect is due to differences in the CETP-mediated HDL remodeling through homotypic transfer of cholesteryl esters and may play an important role in atherosclerosis as pre-β-HDL are important acceptors for ABCA1-mediated cholesterol efflux. Our genetic study could not distinguish between CETP modulation or inhibition, as measurements of ABCA1-mediated efflux or HDL subtypes were not available. CETP may also have an important intracellular role in storing triglycerides and cholesteryl esters in lipid droplets. ^45,46^. Whether this activity was altered in our genetic models or played a role in the effect modification by BMI remains to be determined. Also, the estimated effects derived from the genetic variants relate to lifelong exposure to lower CETP concentrations, which may differ from the effects of short-term exposure to pharmacological inhibition of CETP. In addition, although the UK Biobank offers large numbers of study participants, the cohort has a limited number of cardiovascular events. A case-control cohort with larger numbers of CAD events may help increase power to assess the translation of the detected modulatory effects of sex and BMI on lipid biomarkers and cholesterol efflux to cardiovascular outcomes. We did not adjust for multiple testing of phenotypes and effect modifiers in this study which evaluated several correlated phenotypes. We reported confidence intervals and provided power analyses to support the interpretation of results. The genetic variants at the CETP gene have concurrent effects on multiple biomarkers, making it difficult to disentangle the effect of the individual biomarkers on cardiovascular outcomes. The polygenic modeling of multivariable exposures may be an interesting approach to consider for this purpose ^47^.

In this study, we have evaluated the effect of a genetically predicted reduction in CETP concentration on lipoproteins, lipid fractions, cholesterol efflux and C-reactive protein. We have found results to be largely concordant with those obtained from clinical trials. Using statistical interaction models, we found that sex and BMI are modifiers of the effect of CETP on lipid biomarkers and cholesterol efflux.

## Data Availability

All UK Biobank data is accessible to health researches following their internal approval process. The process to gain access is described here: https://www.ukbiobank.ac.uk/enable-your-research/apply-for-access

## Acknowledgments

We thank the UK Biobank for providing the data under Application Number 20168. We thank the Montreal Heart Institute (MHI) Biobank for providing access to samples and data. We thank members of the jury of MAL’s doctoral thesis for helpful comments.

