## Supplementary Appendix for "Study of effect modifiers of genetically predicted CETP reduction"

### Effect modification by sex and BMI

In our analyses, both sex and BMI modified how a genetically predicted CETP reduction influenced lipid and lipoprotein levels. There is also evidence that there are sex differences in the interaction between adiposity and lipids hinting at a possible three-way interaction.

The three way interaction term including sex, BMI and genetic CETP reduction was significant for LDL-c levels ($p = 4.1 \times 10^{-4}$) and had concordant effects on apoB levels (p = 0.001). As seen on the marginal effect plots (Supplementary Figure 10), BMI influences the effect of CETP on LDL-c and apoB in men with higher BMI values associated with a smaller decrease in LDL-c. This pattern is not seen in women and BMI does not influence the reduction of LDL-c with genetically lower CETP concentration. We did not detect a significant three-way interaction (sex, BMI, genetic CETP reduction) for cardiovascular outcomes, but statistical power was likely limited. In the analysis based on rs1800775, results for biomarkers were similar to those obtained with the score (Supplementary Figure 11).

We also estimated how sex and BMI may modifiy the effect of a genetically lower CETP on cholesterol efflux in the MHI Biobank. There was evidence for a three-way interaction between sex, BMI and the CETP score on basal efflux (p = 0.037) and stimulated efflux (p = 0.007). The marginal effects in men and women with fixed BMI are presented in Supplementary Figure 12. In men, increasing BMI may increase the CETP associated cAMP-stimulated cholesterol efflux (subgroup CETP by BMI interaction p = 0.11) but not its effect on basal efflux (p = 0.53), whereas in women increasing BMI reduced the effect of the CETP score on both stimulated (p = 0.045) and basal efflux (p = 0.050).

For cardiovascular events, the three-way interaction p-values with the genetic score were 0.43 for revascularization procedures, 0.062 for MI, 0.058 for CAD (“soft” definition) and 0.15 for CAD (“hard” definition). In results based on rs1800775, the direction of the interaction in men was inconsistent with the observed effects on biomarkers (increasing BMI conferred a stronger protective effect for all cardiovascular endpoints). The slope of the effect modification of CETP by BMI was inverted in women compared to men for cardiovascular endpoints.

### Results from power analyses

#### *Effect modification by sex*

We conducted power analyses to assess our limit of detection based on the sample size and the prevalence of CAD in men and women in the UK Biobank dataset. For the association of one s.d. reduction in the CETP genetic score with CAD, we calculated that for an OR of 0.98 in men an OR of 0.95 or less in women was sufficient to reach 80% power to detect a significant interaction effect between the CETP genetic score and sex (Supplementary Figure 3).

#### *Effect modification by BMI*

Using simulations, we estimated the smallest detectable effect modification by BMI of the association between genetically-predicted CETP levels and cardiovascular outcomes (Supplementary Figure 8) at 80% power to be $\beta_{itx}$ = 0.015. This represents an OR per s.d. decrease in the CETP score of 0.96 for the normal BMI range versus 0.99 for obese individuals. In our analyses, the product term between the CETP score and BMI was 0.010 (95% CI -0.00042, 0.020) and we estimate our power to detect a statistically significant effect of this magnitude at 48%.
