## Supplementary Figures and Tables for "Study of effect modifiers of genetically predicted CETP reduction"

### Supplementary Tables and Figures

**Supplementary Table 1**. *Correlation coefficient between the different CETP genetic scores or the rs1800775 variant. The upper triangular matrix shows the Pearson correlation coefficient and the lower triangular shows the squared coefficients. All scores were calculated using the p-value thresholding and LD clumping approach, except for the “Blauw et al. condi. indep.” score which is based on the conditionally independent variants reported in the original study.*

| | **MAGNETIC HDL diameter** | **MAGNETIC L-HDL-CE** | **MAGNETIC S-VLDL-TG** | **Blauw *et al.* GRS** | **Blauw *et al.* condi. Indep.** | **SNP rs1800775** |
|---|---|---|---|---|---|---|
| **MAGNETIC HDL diameter** | 1 | 1.00 | -1.00 | -0.87 | -0.94 | 0.72 |
| **MAGNETIC L-HDL-CE** | 1.00 | 1 | -1.00 | -0.87 | -0.94 | 0.72 |
| **MAGNETIC S-VLDL-TG** | 0.99 | 0.99 | 1 | 0.87 | 0.93 | -0.71 |
| **Blauw *et al.* GRS** | 0.76 | 0.76 | 0.76 | 1 | 0.87 | -0.71 |
| **Blauw *et al.* condi. Indep.** | 0.89 | 0.89 | 0.86 | 0.75 | 1 | -0.74 |
| **SNP rs1800775** | 0.51 | 0.51 | 0.51 | 0.51 | 0.55 | 1 |

**Supplementary Table 2**. *Codes used to define cardiovascular endpoint events in the UK Biobank. Events from either the hospitalization or death records were used and the procedure codes (OPCS codes) are from the hospitalization records.*

| **Variable** | **Definition** |
|---|---|
| Myocardial infarction (MI) | ICD10 codes: I21, I22, I23, I25.2 or ICD9 codes: 410, 412, 411.0, 429.79 |
| Percutaneous coronary intervention or coronary artery bypass graft (PCI/CABG) | OPCS codes: K40, K41, K42, K43, K44, K45, K46, K49, K50, K75 |
| Coronary artery disease (CAD) - soft | PCI/CABG or ICD9 codes 410-414 (except for aneurysms: 414.1) or ICD10 codes I20-I25 |
| Coronary artery disease (CAD) - hard | Combination of MI, PCI/CABG or hospitalization or death for unstable angina coded a I20.0 as the primary cause of death or hospitalization. For this variable, cases for the “soft” CAD definition that would be controls for the “hard” CAD definition are set as missing so that they are excluded from the analyses. |

**Supplementary Table 3**. *Effect per alternative allele of rs1800775 (CETP -629C>A) on biomarkers and cardiovascular events. Coefficients for continuous variables are from a linear regression model adjusted for age, sex and the first 10 principal components and are expressed in standard deviation of the outcome per “A” allele of the rs1800775 SNP. Odds ratios for cardiovascular outcomes are estimated using a logistic regression model adjusted for the same covariates.*

| **Outcome** | **n or n cases** | **Coefficient or odds ratio (95% CI)** | **p-value** |
|---|---|---|---|
| **Biomarkers** | | | |

| | | | |
|---|---|---|---|
| lipoprotein(a) | 315,214 | -0.0143 (-0.0182, -0.0106) | $1.4 \times 10^{-13}$ |
| C-reactive protein | 394,165 | 0.0041 (-0.0003, 0.0084) | 0.070 |
| HDL cholesterol | 362,468 | 0.198 (0.194, 0.203) | $<10^{-300}$ |
| Apolipoprotein A | 360,451 | 0.156 (0.151, 0.160) | $<10^{-300}$ |
| LDL cholesterol | 394,287 | -0.028 (-0.033, -0.024) | $3.2 \times 10^{-36}$ |
| Apolipoprotein B | 393,089 | -0.041 (-0.045, -0.037) | $1.4 \times 10^{-73}$ |
| Basal cholesterol efflux (MHI Biobank) | 5,213 | 0.120 (0.0835, 0.156) | $1.1 \times 10^{-10}$ |
| cAMP-stimulated cholesterol efflux (MHI Biobank) | 5,212 | 0.0919 (0.0547, 0.129) | $1.3 \times 10^{-6}$ |
| **Cardiovascular outcomes** | | | |
| Coronary artery disease ("soft") | 44,713 | 0.973 (0.959, 0.987) | 0.00015 |
| Coronary artery disease ("hard") | 26,342 | 0.968 (0.950, 0.986) | 0.00047 |
| Myocardial infarction | 18,559 | 0.982 (0.962, 1.000) | 0.10 |
| Percutaneous coronary intervention or coronary artery bypass graft | 16,941 | 0.970 (0.948, 0.992) | 0.0068 |

**Supplementary Table 4**. *Drug codings to define statin users in the UK Biobank.*

| UK biobank Data-Coding 4 | Prescription |
|---|---|
| 1140861958 | simvastatin |
| 1141146234 | atorvastatin |
| 1141146138 | lipitor 10mg tablet |
| 1141192410 | rosuvastatin |
| 1141192736 | ezetimibe |
| 1140888648 | pravastatin |
| 1141188146 | simvador 10mg tablet |
| 1141192414 | crestor 10mg tablet |
| 1140881748 | zocor 10mg tablet |
| 1141200040 | zocor heart-pro 10mg tablet |

**Supplementary Table 5**. Interaction between the CETP genetic score and **sex** on the additive scale (RERI and interaction contrast) based on the logistic regression model in the UK Biobank.

| Outcome | Odds ratio RERI (95% CI) | Interaction contrast (bootstrap 95% CI) |
|---|---|---|
| Coronary artery disease (“soft”) | 0.031 (-0.006, 0.070) | 0.0011 (-0.00074, 0.0029) |
| Coronary artery disease (“hard”) | **0.082 (0.020, 0.146)** | **0.0017 (0.00024, 0.0032)** |
| Myocardial infarction | 0.050 (-0.018, 0.121) | 0.00078 (-0.00042, 0.0019) |
| Percutaneous coronary intervention or coronary artery bypass graft | 0.091 (-0.003, 0.188) | 0.00099 (-0.00012, 0.0021) |

**Supplementary Table 6**. ANOVA results for the nonlinear interaction models of the CETP genetic score and BMI on biomarkers and cardiovascular outcomes. Linear regression was used for biomarkers and logistic regression was used for cardiovascular outcomes. The presented association statistics are F statistics for the former and $\chi^2$ for the latter. Both the CETP score and BMI are modeled using restricted cubic splines with 4 knots.

| Outcome | Factor | Statistic (F or chi2) | d.f. | P-value |
|---|---|---|---|---|
| *Biomarkers (modeled using linear regression)* | | | | |
| Lipoprotein(a)* | All interactions (BMI and CETP) | 2.14 | 9 | 0.0233 |
| | BMI | 15.55 | 12 | < 0.0001 |
| | Nonlinear effects | 22.09 | 8 | < 0.0001 |
| | CETP genetic score | 6.91 | 12 | < 0.0001 |
| | Nonlinear effects | 0.94 | 8 | 0.4793 |
| C-reactive protein | All interactions (BMI and CETP) | 0.89 | 9 | 0.5299 |
| | BMI | 7662.84 | 12 | < 0.0001 |
| | Nonlinear effects | 19.81 | 8 | < 0.0001 |
| | CETP genetic score | 1.47 | 12 | 0.1265 |
| | Nonlinear effects | 0.66 | 8 | 0.7247 |
| HDL cholesterol | All interactions (BMI and CETP) | 37.41 | 9 | < 0.0001 |
| | BMI | 4975.76 | 12 | < 0.0001 |
| | Nonlinear effects | 189.61 | 8 | < 0.0001 |
| | CETP genetic score | 1242.72 | 12 | < 0.0001 |
| | Nonlinear effects | 8.51 | 8 | < 0.0001 |
| Apolipoprotein A | All interactions (BMI and CETP) | 11.72 | 9 | < 0.0001 |
| | BMI | 2632.38 | 12 | < 0.0001 |
| | Nonlinear effects | 77.75 | 8 | < 0.0001 |
| | CETP genetic score | 683.50 | 12 | < 0.0001 |
| | Nonlinear effects | 2.05 | 8 | 0.0365 |
| LDL cholesterol | All interactions (BMI and CETP) | 3.68 | 9 | 0.0001 |

| | | | | |
|---|---|---|---|---|
| | BMI | 521.46 | 12 | < 0.0001 |
| | Nonlinear effects | 704.12 | 8 | < 0.0001 |
| | CETP genetic score | 20.57 | 12 | < 0.0001 |
| | Nonlinear effects | 3.17 | 8 | 0.0013 |
| Apolipoprotein B | All interactions (BMI and CETP) | 2.29 | 9 | 0.0147 |
| | BMI | 733.80 | 12 | < 0.0001 |
| | Nonlinear effects | 638.04 | 8 | < 0.0001 |
| | CETP genetic score | 38.91 | 12 | < 0.0001 |
| | Nonlinear effects | 2.70 | 8 | 0.0058 |
| Basal cholesterol efflux (MHI Biobank) | All interactions (BMI and CETP) | 0.66 | 9 | 0.7473 |
| | BMI | 17.55 | 12 | < 0.0001 |
| | Nonlinear effects | 1.91 | 8 | 0.0537 |
| | CETP genetic score | 5.84 | 12 | < 0.0001 |
| | Nonlinear effects | 0.64 | 8 | 0.7437 |
| cAMP-stimulated cholesterol efflux (MHI Biobank) | All interactions (BMI and CETP) | 0.49 | 9 | 0.8815 |
| | BMI | 4.27 | 12 | < 0.0001 |
| | Nonlinear effects | 0.50 | 8 | 0.8537 |
| | CETP genetic score | 3.67 | 12 | < 0.0001 |
| | Nonlinear effects | 0.64 | 8 | 0.7439 |
| Cardiovascular outcomes *(modeled using logistic regression)* | | | | |
| Coronary artery disease ("soft") | All interactions (BMI and CETP) | 8.10 | 9 | 0.5244 |
| | BMI | 4892.66 | 12 | < 0.0001 |
| | Nonlinear effects | 128.51 | 8 | < 0.0001 |
| | CETP genetic score | 30.67 | 12 | 0.0022 |
| | Nonlinear effects | 4.84 | 8 | 0.7742 |
| Coronary artery disease ("hard") | All interactions (BMI and CETP) | 6.16 | 9 | 0.72 |
| | BMI | 2798.71 | 12 | < 0.0001 |
| | Nonlinear effects | 68.01 | 8 | < 0.0001 |
| | CETP genetic score | 25.36 | 12 | 0.0132 |
| | Nonlinear effects | 3.19 | 8 | 0.9219 |
| Myocardial infarction | All interactions (BMI and CETP) | 8.64 | 9 | 0.4716 |
| | BMI | 1857.02 | 12 | < 0.0001 |
| | Nonlinear effects | 69.72 | 8 | < 0.0001 |
| | CETP genetic score | 15.49 | 12 | 0.2157 |
| | Nonlinear effects | 5.71 | 8 | 0.6798 |

| | | | | |
|---|---|---|---|---|
| Percutaneous coronary intervention or coronary artery bypass graft | All interactions (BMI and CETP) | 5.49 | 9 | 0.7893 |
| | BMI | 1254.46 | 12 | < 0.0001 |
| | Nonlinear effects | 122.46 | 8 | < 0.0001 |
| | CETP genetic score | 22.25 | 12 | 0.0349 |
| | Nonlinear effects | 3.37 | 8 | 1 |

* The Lp(a) genetic score was added as a covariate.

**Supplementary Table 7**. Interaction between the CETP genetic score and **body mass index** on the additive scale (RERI and interaction contrast) based on the logistic regression model in the UK Biobank.

| Outcome | Odds ratio RERI (95% CI) | Interaction contrast (bootstrap 95% CI) |
|---|---|---|
| Coronary artery disease (“soft”) | -0.002 (-0.017, 0.012) | -0.00030 (-0.0013, 0.00076) |
| Coronary artery disease (“hard”) | -0.001 (-0.020, 0.018) | $-9.5 \times 10^{-5}$ (-0.00090, 0.00068) |
| Myocardial infarction | -0.005 (-0.025, 0.016) | -0.00014 (-0.00073, 0.00043) |
| Percutaneous coronary intervention or coronary artery bypass graft | 0.00039 (-0.021, 0.022) | $-5.8 \times 10^{-6}$ (-0.00051, 0.00051) |

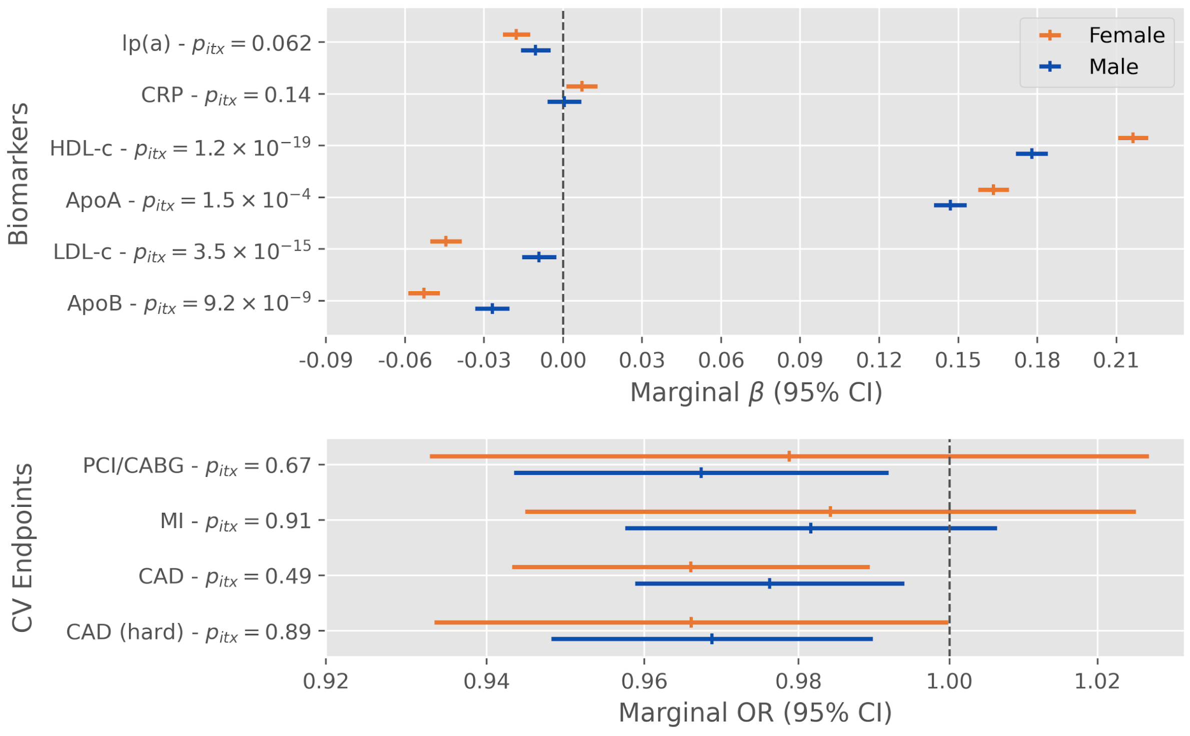


**Supplementary Figure 1.** *Effect modification of* ***rs1800775*** *(CETP -629C>A) alternative alleles by sex on biomarkers and cardiovascular endpoints in the UK Biobank. Displayed*

*p-values ($p_{itx}$) are for the two-sided test of the product term between the SNP and a binary sex indicator variable.*

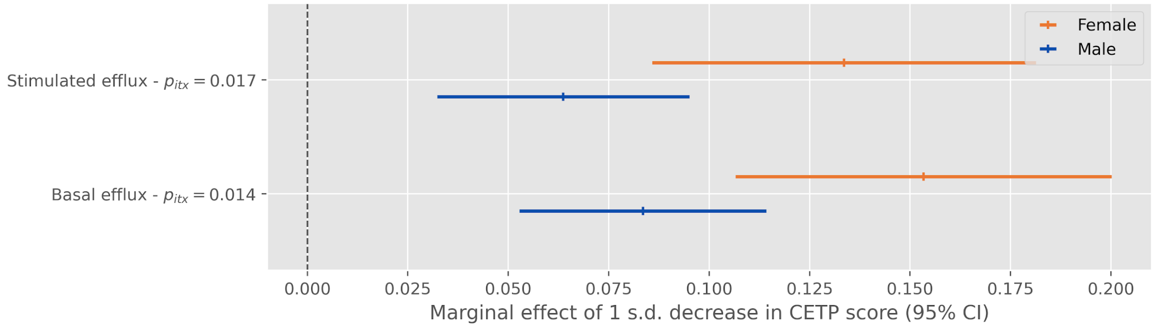


**Supplementary Figure 2**. *Effect modification of a 1 standard deviation decrease in the CETP concentration genetic score by sex on cholesterol efflux in the MHI Biobank. Displayed p-values ($p_{itx}$) are for the two-sided test of the product term between the CETP score and a binary sex indicator variable.*

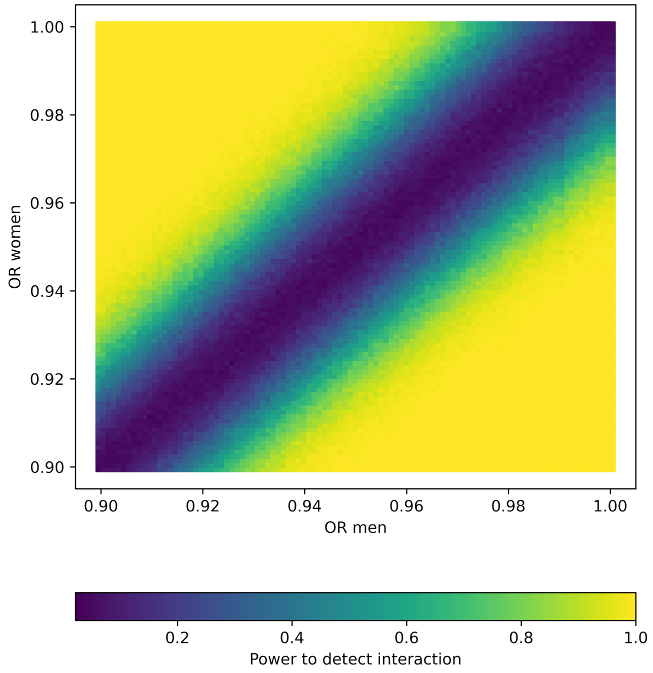


**Supplementary Figure 3**. *Power to detect a difference between men and women in the association between a standardized genetic score and a coronary artery disease. The results of this simulation are for 500 simulated datasets for different combinations of OR in men and*

*women and given the sample size (n=413,138) and prevalence of coronary artery disease ("soft" definition, 10.8%) in our dataset.*

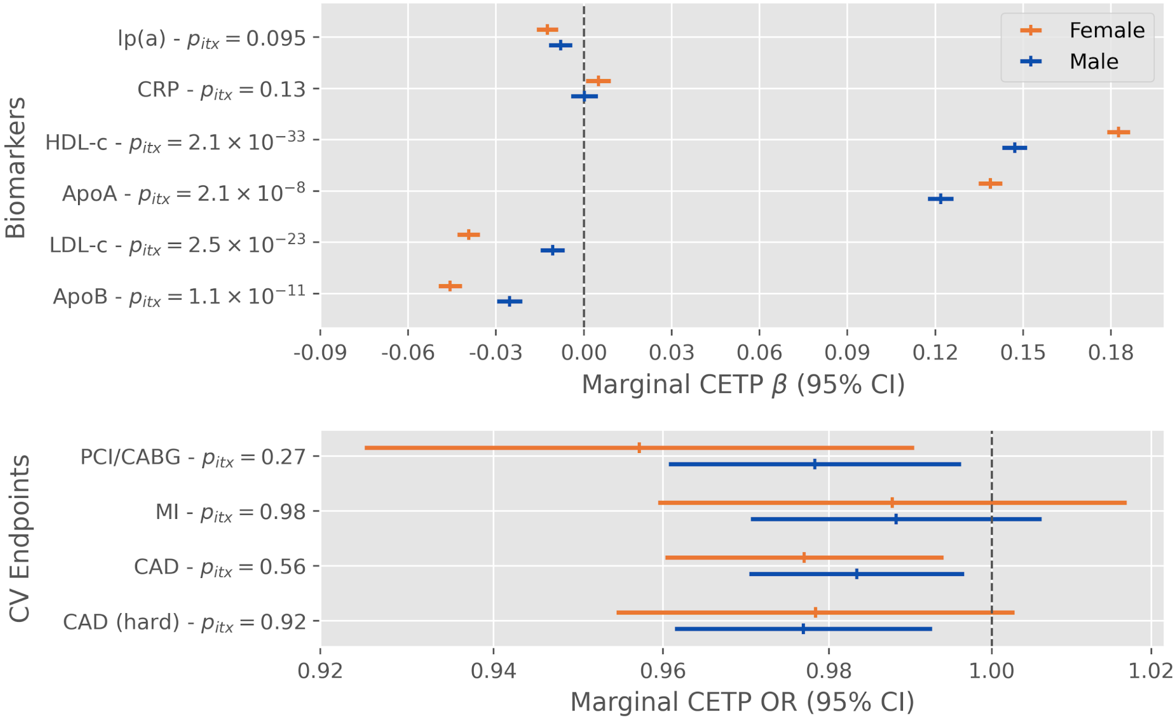


**Supplementary Figure 4**. *Effect modification of a 1 standard deviation decrease in the CETP concentration genetic score by sex on biomarkers and cardiovascular endpoints* ***adjusting for self-reported statin use*** *in the UK Biobank. Displayed p-values ($p_{itx}$) are for the two-sided test of the product term between the CETP score and a binary sex indicator variable.*

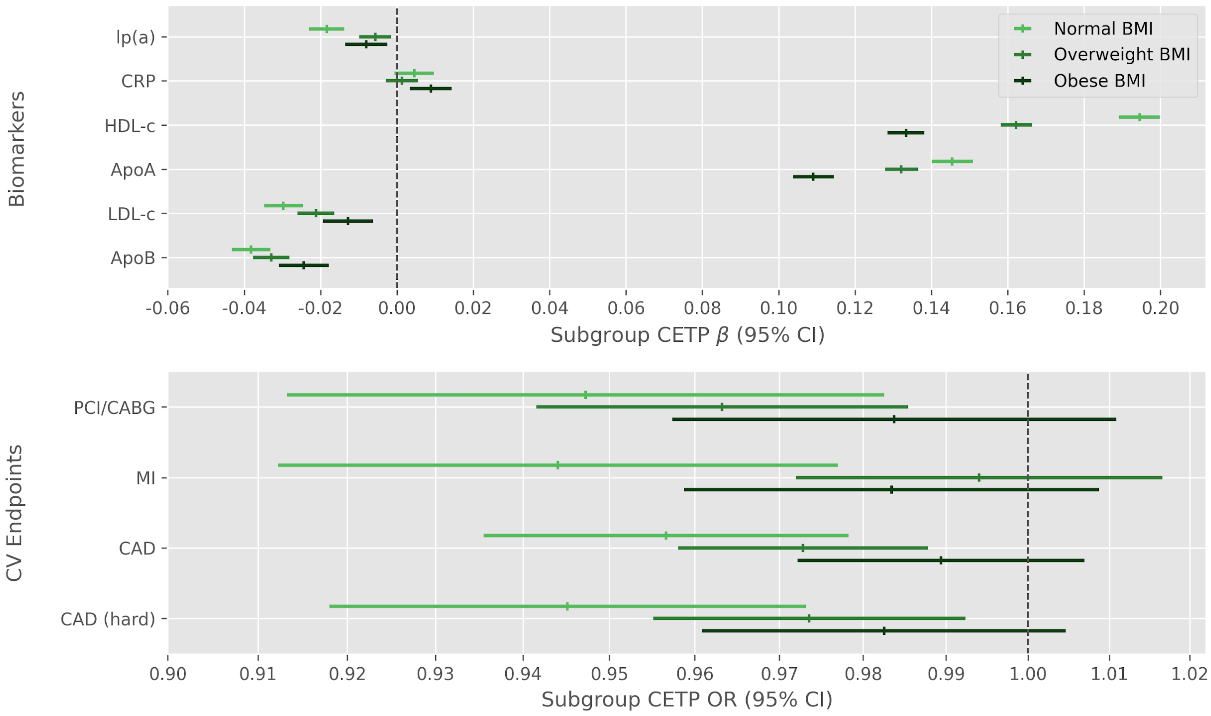


**Supplementary Figure 5.** *Subgroup effect of a 1 standard deviation decrease in the CETP concentration genetic score on biomarkers and cardiovascular endpoints by BMI class in the UK Biobank.*

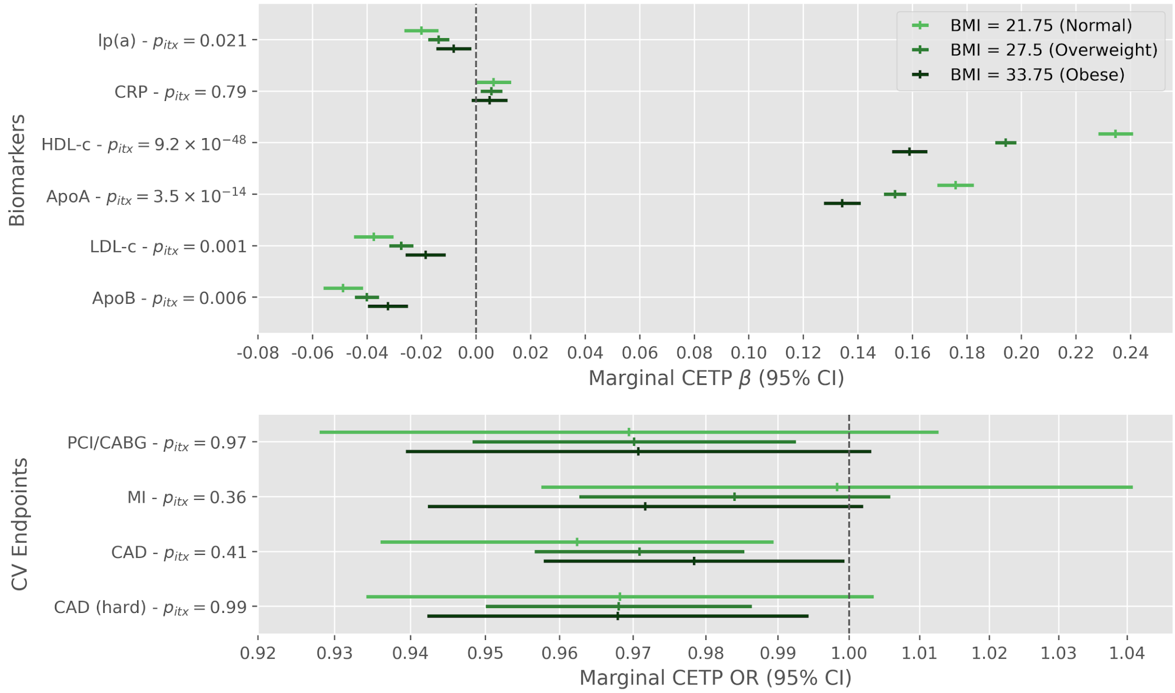


**Supplementary Figure 6.** *Effect modification of rs1800775 (CETP -629C>A) alternative alleles by BMI on biomarkers and cardiovascular endpoints in the UK Biobank. Displayed*

*p-values ($p_{itx}$) are for the two-sided test of the product term between the SNP and standardized body mass index.*

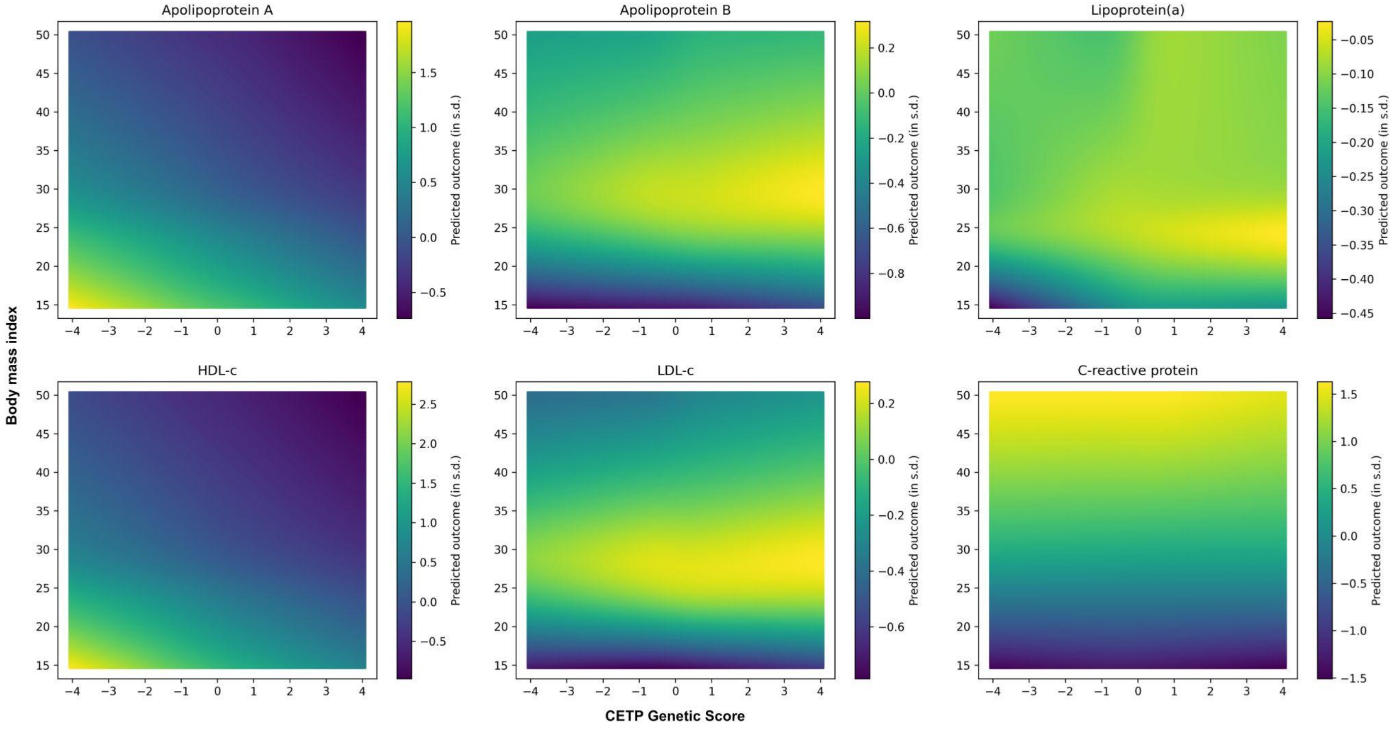


**Supplementary Figure 7**. *Expected value of the standardized biomarkers predicted by linear regression models including interacting splines for the BMI and the genetic score. The CETP genetic score was set to values between -4 and 4 (x axis) and the BMI was set to values between 15 and 50 (y axis).*

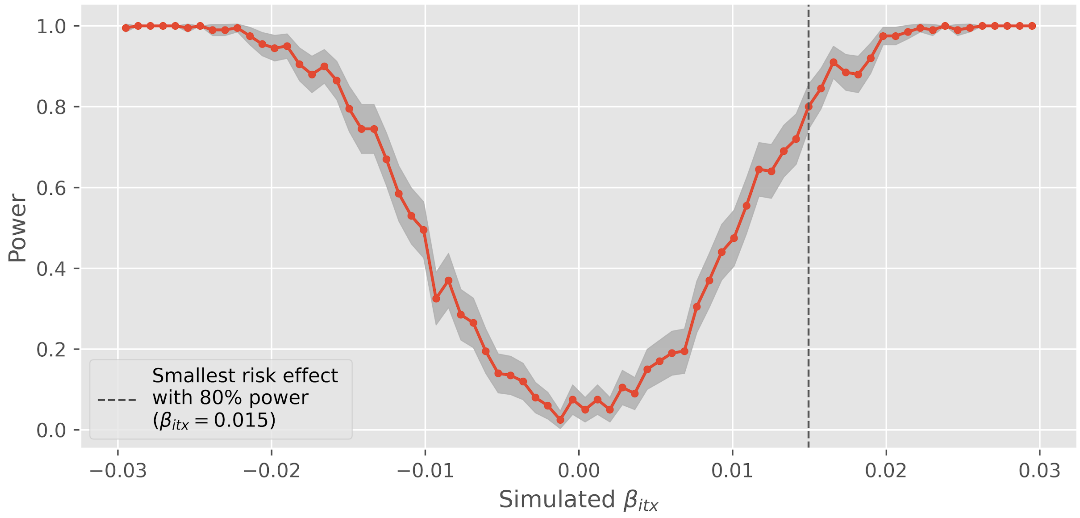


**Supplementary Figure 8**. *Simulation-based power analysis to detect an additive interaction effect between a continuous CETP score and standardized BMI. This simulation sets as static*

*parameters the prevalence of CAD and the mean effect of a reduction in the CETP score. Results are based on 200 simulation replicates of 413,138 individuals.*

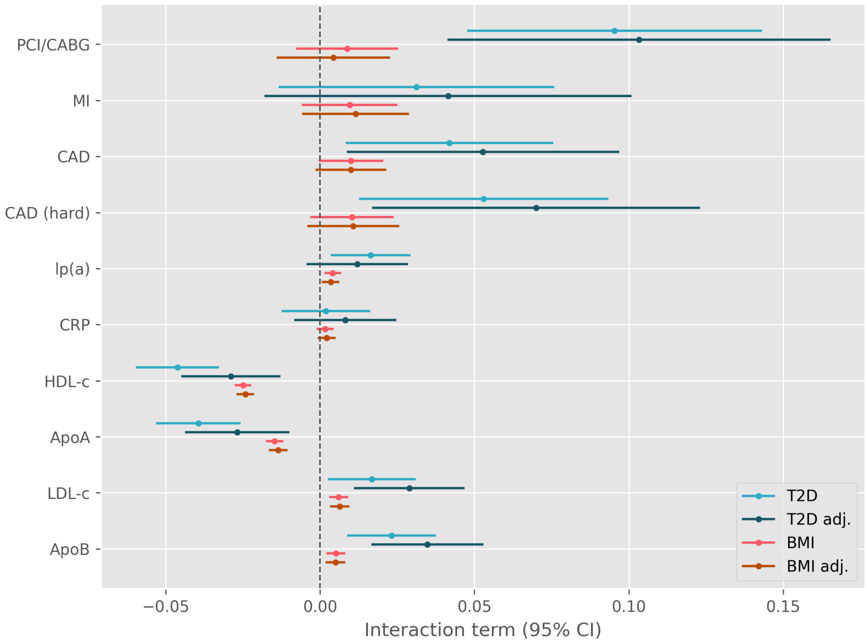


**Supplementary Figure 9.** *Interaction coefficients between the CETP genetic score and type II diabetes and BMI with and without adjustment for the other variable in the UK Biobank. The unadjusted model includes the product interaction term between CETP and type II diabetes or BMI whereas the adjusted model includes the three-way product interaction.*

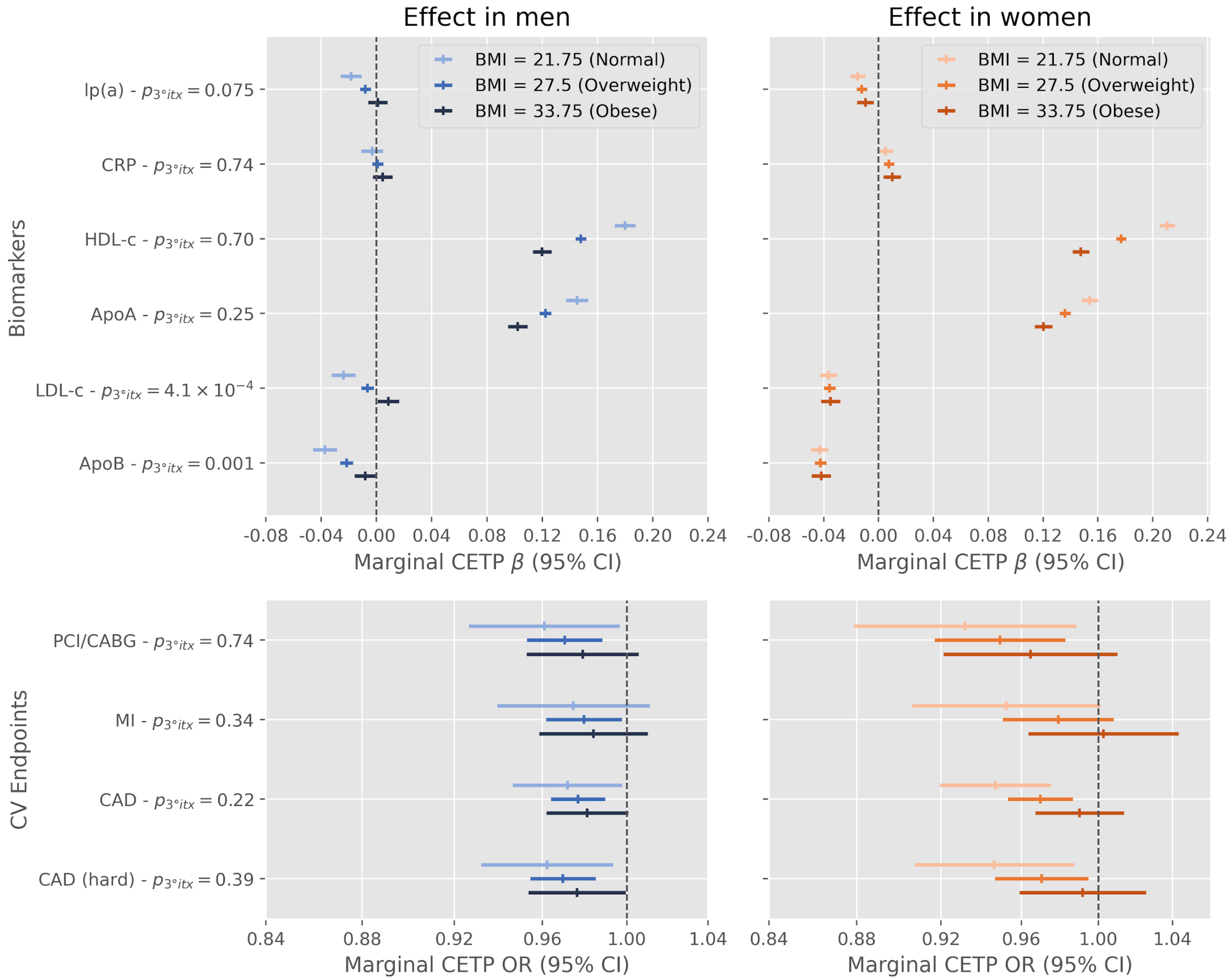


**Supplementary Figure 10.** *Effect modification of a 1 standard deviation decrease in the CETP concentration genetic score by BMI and sex on biomarkers and cardiovascular endpoints in the UK Biobank. Displayed p-values ($p_{3°itx}$) are for the two-sided test of the three way product term between the CETP score, standardized body mass index and a binary variable for sex.*

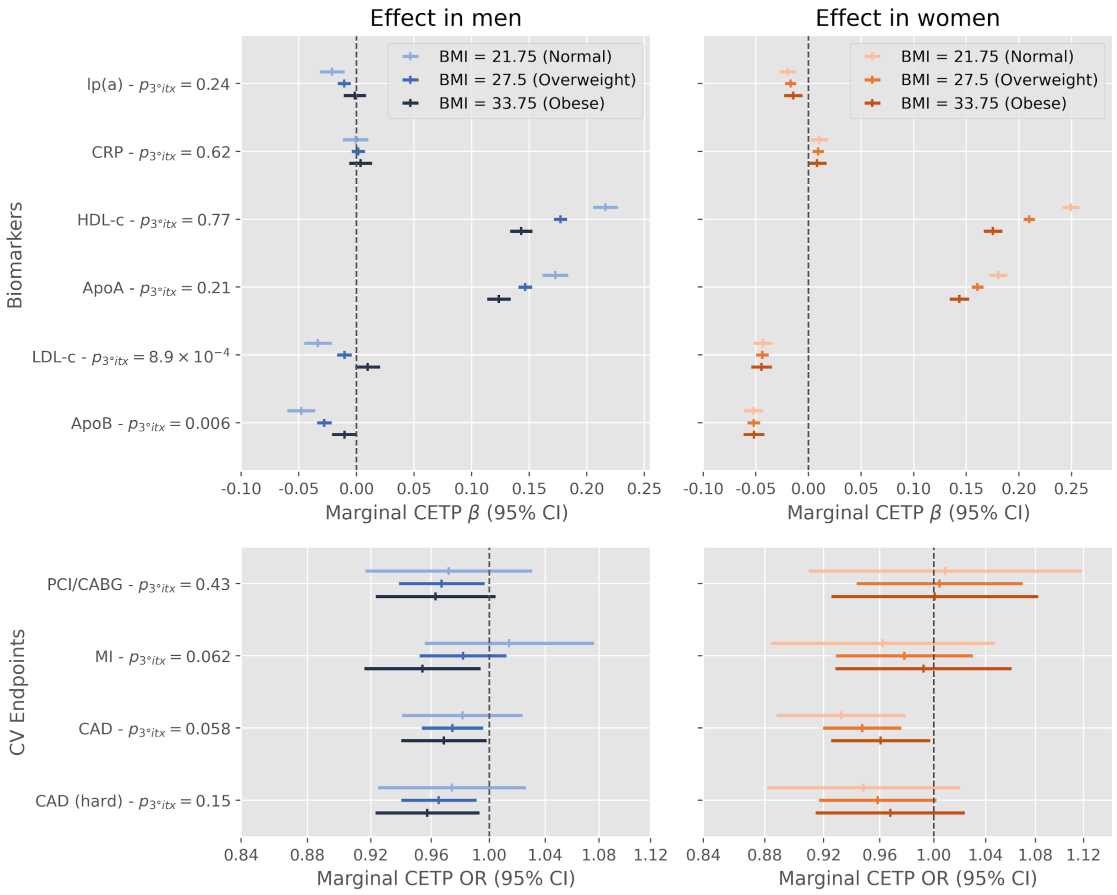


**Supplementary Figure 11.** *Effect modification of rs1800775 (CETP -629C>A) alternative alleles by BMI and sex on biomarkers and cardiovascular endpoints in the UK Biobank. Displayed p-values ($p_{3°itx}$) are for the two-sided test of the three-way product term between the SNP, a binary variable for sex and standardized body mass index.*

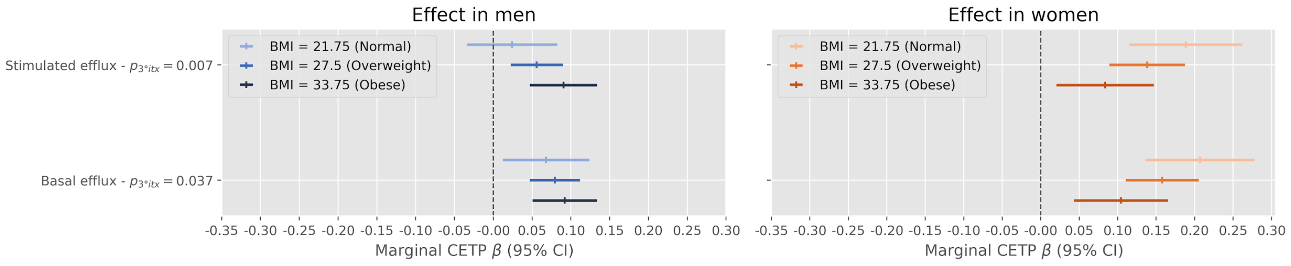


**Supplementary Figure 12** *Effect modification of the CETP score on cholesterol efflux in the MHI Biobank by sex and BMI.*
