## Supplementary Methods for "Study of effect modifiers of genetically predicted CETP reduction"

#### 1 UK Biobank

To assess the effect of genetic CETP modulation on biomarkers and cardiovascular diseases by sex and BMI, we used the UK Biobank resource. The UK Biobank is a population-based longitudinal cohort of more than 500,000 individuals [1]. At recruitment, a touchscreen-based questionnaire followed-up by a verbal interview with a nurse was used to assess self-reported diseases and medications. Linkage with hospitalization and death registries also allows acute events such as cardiovascular events to be well captured. Anthropomorphic measurements were also taken and an extensive panel of blood and urine biomarkers captures many laboratory measurements including major lipoprotein fractions. Finally, genetic data based on a genotyping array and imputation is available on all participants.

Biochemical markers were measured from samples of urine, packed red blood cells and serum collected at baseline for all participants. For logistics reasons, the UK Biobank opted for centralized processing of samples in a high throughput facility. Participants were not required to fast before the collection of biological samples.

For the analysis of blood biomarkers including lipoproteins and cholesterol levels, we transformed the variables as needed to obtain an approximately normal distribution and then standardized the values (units are reported in the main text Table 1). Because we included different biomarkers in our analyses, standardization allows unified reporting of effects in units of standard deviation. For all continuous measurements, we used values from the baseline visit and we used the mean if multiple measurements were available.

To define type 2 diabetes, we opted to rely on the self-reported diseases from the verbal interview with a nurse. We extracted all individuals who self-reported "diabetes" (coded as 1220 in variable #20002) or its children variables ("gestational diabetes" coded 1221, "type 1 diabetes" coded 1222, "type 2 diabetes" coded 1223 or "diabetes insipidus" coded 1521). Then, we excluded from the analyses individuals reporting "type 1 diabetes" or reporting "gestational diabetes" or "diabetes insipidus" with no record of "type 2 diabetes". The remaining individuals were assumed to have type 2 diabetes and individuals that did not report any of the preceding codes were used as controls.

For cardiovascular events, preliminary analyses revealed that self-reported data could lack precision and so we relied on ICD10 codes from hospitalization or death records. Revascularization procedures (percutaneous coronary interventions [PCI] and coronary artery bypass graft [CABG]) were defined using OPCS procedure codes in the linkage with hospitalization data. The specific codes used to define the events (based on hospitalizations, deaths or procedures) are described in Supplementary Table 3.

For the statin use status, we extracted all the self-reported medications (coded 20003). All drugs classified under the ATC code C10B were used to define statin users. We used the UK Biobank Self Reported Medication Data parsing and matching software (available `https://github.com/PhilAppleby/ukbb-srmed/`) to achieve this coding. This procedure resulted in the identification of 83,385 statin users based on 10 drugs. The specific codes used to define this composite are described in Supplementary Table 4.

#### 1.1 Genetic quality control

The genetic quality control steps used to avoid risk of confounding due to ethnicity, relatedness or incorrect genotyping has been previously described [2].

Briefly, we excluded variants or individuals with a missing rate above 2%. We compared the genetic and self-reported sex variables to validate sample matching between the genetic and phenotypic data. Individuals with discrepencies or aneuploidies were removed from the analysis dataset. To avoid bias due to population stratification, we only selected individuals from the majority population in the UK Biobank. We excluded individuals not self-reporting as of European descent or falling outside of a manually defined region based on the principal component analysis projections. Related individuals were also excluded from analysis using a kinship coefficient of 0.0884 as the cutoff (corresponding to a 3rd degree relationship). After genetic quality control, a total of 413,138 individuals remained.

### 2 Montreal Heart Institute Biobank

Participants to the Montreal Heart Institute (MHI) Biobank and hospital cohort were recruited from different MHI departments and its affiliated prevention centre (EPIC) between 2006 and 2016. Blood, DNA, and plasma are collected at baseline and stored at the Pharmacogenomics Centre at MHI. All MHI Biobank participants provided informed consent and the study was approved by the MHI scientific and ethics review committees.

#### 2.1 Cholesterol efflux measurements

Basal and cAMP-stimulated cholesterol efflux were measured using plasma from a subset of 5,215 participants of the MHI Biobank. The procedure for the efflux measurements was previously published elsewhere [3]. Briefly, plasma was obtained from venous blood samples collected on potassium-EDTA coated tubes (BD Vacutainers), centrifuged as per the manufacturer's protocol and frozen at -80°C until analysis. Samples were then thawed at 4°C and the cholesterol efflux capacity was measured in vitro with J774 macrophages. Cells were grown for 24 hours in presence of tritiated ($^3H$) cholesterol (2 $\mu Ci/ml$). After an 18 hours equilibration period, patient plasma depleted of apoB-containing lipoproteins with PEG6000 or control serum was added in triplicate wells for 4 hours. The concentration measurement was selected from dose-response curves obtained from pooled human plasma to avoid saturation of the efflux signal and to allow detection of changes in efflux capacity in any direction. Aliquots of cell-free culture medium and J774 cell homogenates were used to measure $^3H$ cholesterol counts with a beta counter (Tricarb, Perkin-Elmer). Cholesterol efflux capacity was defined as the ratio of $^3H$-cholesterol found in the medium to the total cholesterol label in each well. Sample batches were tested in parallel with the same pool of normolipidemic human serum to calculate the sample/control ratio. We rejected cholesterol efflux capacity measurements that were outside of the historical mean $\pm$ 2 standard deviations.

To avoid batch effects in the statistical analysis, we used the residuals of the multivariable regression of efflux measurements on a factor variable representing the day of analysis. The residuals were subsequently standardized and used as the dependant variable in our analyses.

### 3 Simulation-based power analyses

#### Simulation for an interaction with body mass index

We are interested in assessing whether BMI modifies the effect of genetically-predicted reduction in CETP concentration on coronary artery disease (CAD). To estimate statistical power to detect such an effect, we simulate interaction terms while fixing the overall disease prevalence, the allele frequencies and marginal effect of the CETP genetic score to observed values from the data. We repeat the simulation and estimate the fraction of simulated datasets where the null hypothesis of $\beta_{itx} = 0$ is rejected at $\alpha = 0.05$ using a conventional logistic regression. The simulation parameters are described below.

| Parameter | Description | Value |
|---|---|---|
| $n$ | Number of individuals | 413,138 |
| $P(Y=1)$ | Prevalence of coronary artery disease in the overall population | 10.8% |
| $Y$ | Coronary artery disease (CAD), the outcome. | Simulated as described in the "Simulation" section |
| $X$ | The CETP genetic score expressed so that 1 unit of X corresponds to a 1 standard deviation decrease in the CETP concentration score (negative of the standardized score of CETP concentration). | Simulated $\mathcal{N}(0,1)$ |
| BMI | The body mass index | Simulated $\mathcal{N}(0,1)$ |
| $\beta_0$ | Intercept for the logistic regression model. Corresponds to the logistic of the prevalence when other covariables are 0. | See section "Simulation model" |
| $\beta_x$ | Marginal effect of a 1 unit increase in $X$ (CETP) on $Y$ (coronary artery disease). In our analyses, a 1 unit increase in $X$ represents a 1 standard deviation reduction in the genetic score of CETP concentration. This is the $\beta_x$ as estimated in a model with no interaction terms. | -0.0255 |
| $\beta_b$ | Coefficient of BMI (direct effect on $Y$). This value is the observational effect of BMI on $Y$ as estimated in the UK Biobank. We used the estimate from a multivariable logistic regression model including age, sex and principal components as covariates. | 0.366 |
| $\beta_{itx}$ | The additive effect modification of $\beta_x$ per unit increase in BMI (*i.e.* product interaction term between BMI and $X$). | Simulated values between -0.03 and 0.03 |

The association model is the following logistic regression model:

$$\ln\left(\frac{P(Y=1)}{1-P(Y=1)}\right) = \beta_0 + \beta_x \cdot X + \beta_b \cdot \text{BMI} + \beta_{itx} \cdot X \cdot \text{BMI} \tag{1}$$

Where the parameters are as previously defined.

#### Simulation

When simulating the outcome (CAD), we want to control for the prevalence ($P(CAD=1)$) and obtain the desired regression coefficients. To achieve this, we use a latent variable model of the logistic regression.

$$Y^* = \beta_0 + \beta_x \cdot X + \beta_b \cdot \text{BMI} + \beta_{itx} \cdot X \cdot \text{BMI} + \epsilon \tag{2}$$

With

$$\begin{aligned} \epsilon &\sim \mathcal{L}(0,1) \\ X &\sim \mathcal{N}(0,1) \\ \text{BMI} &\sim \mathcal{N}(0,1) \end{aligned}$$

and the coefficients set as previously described except for $\beta_0$ which we will discuss later. The resulting continuous latent variable ($Y^*$) is used to simulate the outcome as follows:

$$Y = \begin{cases} 1 & \text{if } Y^* > 0 \\ 0 & \text{otherwise} \end{cases}$$

We then use $\beta_0$ to control the prevalence (*i.e.* $P(Y = 1)$) as $Y^*$ is positive if and only if

$$-\beta_0 < \beta_x \cdot X + \beta_b \cdot \text{BMI} + \beta_{itx} \cdot X \cdot \text{BMI} + \epsilon$$

Using the normal approximation for the right hand side of the former, we have:

$$\begin{aligned} &Z = \beta_x \cdot X + \beta_b \cdot \text{BMI} + \beta_{itx} \cdot X \cdot \text{BMI} + \epsilon \\ &Z \sim \mathcal{N}\left(0, Var\left(\beta_x X + \beta_b \text{BMI} + \beta_{itx} \cdot X \cdot \text{BMI} + \epsilon\right)\right) && \text{Normal approximation and } X \text{ standard normal} \\ &Z \sim \mathcal{N}\left(0, \beta_x^2 + \beta_b^2 + Var(\beta_{itx} \cdot X \cdot \text{BMI}) + \frac{\pi^2}{3})\right) && \text{Variance of standard normal and logistic} \\ &Z \sim \mathcal{N}\left(0, \beta_x^2 + \beta_b^2 + \beta_{itx}^2 + \frac{\pi^2}{3}\right) && \text{Assuming X and BMI independant} \end{aligned}$$

And we can use the normal quantile function to determine the value of $\beta_0$ resulting in the observed prevalence.

$$\begin{aligned} P(Y = 1) = p &= P(-\beta_0 < Z) \\ &= 1 - \Phi_Z(-\beta_0) \iff \\ \beta_0 &= -\Phi_Z^{-1}(1 - p) \end{aligned}$$

##### Implementation (R)

We implemented the simulation in R. Power computations are done by simulating an outcome, fitting a logistic regression model and empirically estimating the power as the fraction of all simulation iterations where the null was rejected at $\alpha = 0.05$.

```
# Fixed parameters
n <- 413138
prevalence <- 44713 / n

# Estimated from CAD ~ bmi + age + sex + PCs model
b_bmi <- 0.3659062

# Marginal effect of CETP at mean BMI
b_cetp <- -0.025514967819954

bmi <- rnorm(n)
cetp <- rnorm(n)

b0 <- -qnorm(
    1 - prevalence,
    mean = 0,
    sd = sqrt(b_cetp^2 + b_bmi^2 + b_itx^2 + pi^2 / 3)
)

y_star <- b0 + b_cetp * cetp + b_bmi * bmi + b_itx * cetp * bmi + rlogis(n)
y <- as.numeric(y_star > 0)
```

##### Power estimation

We estimated power by repeatadly simulating datasets with fixed parameters and estimating the proportion of all simulations where the null hypothesis is rejected using the conventional hypothesis testing framework. Specifically, we test the hypothesis that the interaction coefficient is different from zero at an $\alpha = 0.05$ threshold and using the Wald statistic.

The standard error of the estimated power is $\sqrt{\hat{p}(1 - \hat{p})/n}$ where $\hat{p}$ is the proportion of simulated datasets where the null is rejected and $n$ is the number of simulated datasets. When plotting power estimates, the standard error is used to construct the 95% confidence interval.

#### Simulation for an interaction with sex

The simulation model for the effect modification by sex was a little bit different. Because the levels in the simulation were binary, we simulated men and women separately and concatenated the results. The proportion of men and women from our dataset and the prevalence of CAD were fixed at observed values. The sex-specific effect of CETP was then used as the simulation parameter using the same strategy as before to control for the prevalence of CAD (the latent logistic regression model).

### 4 Construction of CETP activity scores

To construct a weighted allele score of CETP activity, we considered various approaches. We constructed a total of 5 scores based on various summary statistics from previous GWAS of plasma CETP concentration [4] or lipid phenotypes measured by nuclear magnetic resonance by the MAGNETIC consortium [5]. In the end, we only selected one representative score for the analyses to avoid redundancy as all the scores we generated were strongly correlated (Supplementary Table 1). All scores were constructed using the LD clumping and p-value thresholding approach based on variants at the CETP locus, except from the score based on the independently associated variants as presented in the original publication of the CETP concentration GWAS. To determine an appropriate p-value threshold for the genetic scores, we estimated the effective number of independent pairs of lipid and variant associations akin to the simpleM correction [6].

Specifically, we estimated the number of independent genetic variants at the CETP locus using a Principal Component Analysis (PCA) of the genotype matrix. Including 50 principal components explained 99% of the variance in CETP genotypes and was used to estimate the effective number of tested variants. Similarly, to estimate the effective number of lipid phenotypes to account for, we used hierarchical clustering of the correlation between variant association coefficients in the MAGNETIC GWAS. We used this dataset to estimate the number of independent phenotypes because it includes an exhaustive number of lipid traits. Manual inspection of the hierarchical clustering dendrogram revealed that there were three major classes of lipids independently affected by CETP. We picked HDL cholesteryl ester, small VLDL triglycerides and large VLDL triglycerides as cluster representatives. Association coefficients for these lipids were thus used when generating candidate scores of CETP activity. Based on the estimates of the effective number of variants and lipids, the p-value threshold was set to $0.05/(3 \times 50) = 3.3 \times 10^{-4}$ for all scores. We fixed the LD threshold at $r^2 = 0.1$ as to not exclude variants correlated by chance and a minimum allele frequency of 1% was used.

### 5 Causal interpretation

Genetic variants have an important property that makes them a good tool for causal inference: they have very few causes. Because they are determined at birth and fixed throughout life, they are not susceptible to reverse causation or environmental confounders, two major sources of bias in observational studies. Mendelian randomization (MR) leverages this advantage of genetic variants to draw causal inferences of the effect of heritable exposures [7, 8].

The current study is similar in spirit to an MR study and we will formalize how the presented associations can be causally interpreted based on the same assumptions as any instrumental variable study. We represented the causal structure of the current experiment as a causal directed acyclic graph (DAG) below: [9]

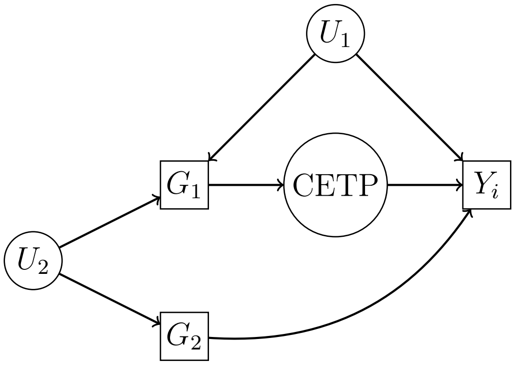


Figure 1: Directed acyclic graph representing the causal structure of the experiment. Squares are used to denote observed variables and circles are used to denote unobserved variables. Arrows represent causal effects such that changes in a parent variable will result in changes in its child.

In this DAG, we can see features that are common in instrumental variable studies such as the classical MR setup. The $G_1$ node represents a genetic predictor of the unobserved CETP activity. In our study, this corresponds to either the CETP concentration score or the CETP -629C>A (rs1800775) variant. The unobserved CETP node represents all aspects of a genetic disruption of CETP in terms of plasma concentration, isoform prevalence, enzymatic activity and substrate preference. In our study this is an unobserved variable that encompasses different aspects of CETP function. The $Y_i$ node represents the outcomes of interest. In our study, it is used to represent alternatively C-reactive protein, lipid fractions, lipoprotein levels, plasma cholesterol efflux capacity or cardiovascular outcomes, captured by the index on the $Y_i$ variable.

Two latent variables ($U_1$ and $U_2$) corresponding to potential confounders are also included. The first variable ($U_1$) is used to represent an effect of population stratification. Specifically, the distribution of $G_1$ is expected to vary between populations simply due to differences in population histories. The distribution of $Y_i$ may also vary across populations because of lifestyle or environmental differences. For example a diet common in one area of the world may influence risk of heart attack in a way that is independent from genetics. If unaccounted for, this common cause of $G_1$ and $Y_i$ (ancestry) will bias the estimate of the effect of $G_1$ on $Y_i$. In our study, we ensured that this unobserved variable was controlled for by only using the largest genetically homogeneous subset of the UK Biobank consisting of individuals of European descent. Additionally, we included principal components capturing population structure to our regression models, which is the conventional way to account for population stratification in genetic association studies.

The second possible confounder is depicted on the graph by the $U_2$ node. This node represents unobserved linkage disequilibrium (LD) inducing a correlation between the $G_1$ variable and a second locus ($G_2$) with a direct effect on $Y_i$. As is conventional in MR, we will assume no direct effect of $G_1$ on $Y$ which includes the effect of the $G_1 \rightarrow U_2 \rightarrow G_2 \rightarrow Y$ path. This assumption also justifies that no arrow from $G_1$ to $Y$ was included in the DAG. In instrumental variable analysis and MR this assumption is called the exclusion restriction. As in any MR study, the causal interpretation of our results requires this unverifiable, but plausible, assumption to hold. Another reason to include a direct effect of $G_1$ on $Y$ is horizontal pleiotropy, a term used to describe a genetic variant with simultaneous effects on distinct pathways. Because the score only includes CETP locus variants associated with plasma concentration, and the rs1800775 "C" allele is known to repress CETP promoter activity and Sp3 transcription factor binding, it is unlikely that the observed effects are through another pathway. In general, studies based on well known variants at a single locus are less likely to suffer from bias due to pleiotropy.

To summarize, we make the following assumptions:

1. **There are no confounders of the $G_1 - Y_i$ relationship (independance).** $U_1$ is accounted for by using a genetically homogeneous subset of the UK Biobank and including principal components in our regression models.
2. $G_1 \rightarrow$ **CETP is non-null (relevance).** This is given by external data validating the effect of rs1800775 on CETP levels and by construction for the score of CETP concentration. We also argue that the effect observed effects on HDL-c levels, a well-known consequence of CETP disruption, supports this assumption.
3. $G_1$ **only affects** $Y_1$ **through CETP (exclusion restriction).** More formally, $G_1 \perp\!\!\!\perp Y_1 \mid \text{CETP}$. We minimized the risk of violating this assumption by only relying on genetic variables known to affect CETP function and located at the CETP locus.

These assumptions are the same as for instrumental variable or MR analyses and allow us to interpret the multivariable regression coefficients described in the main text as the total effect of $G_1$ on $Y_i$. Because this total effect is only mediated by CETP, under our assumptions, it represents the average causal effect of genetic CETP disruption.

In our manuscript, the focus is on effect modifiers of the genetically predicted reduction in CETP on biomarkers and cardiovascular outcomes. The question of effect modification in causal effects is interesting and currently scarcely discussed in the literature, especially in the parametric and multivariable context. Stratification has been suggested as the natural way to identify effect modification and so we reported marginal subgroup effects for all of the considered effect modifiers in the main text [10].

Because the CETP node is unobserved, we are not able to distinguish effect modification in the $G_1 \rightarrow CETP$ from the $CETP \rightarrow Y_i$ effects. This distinction is important and represents a limitation of our study if generalization to pharmacological CETP inhibition is sought. Concretely, if the effect modification only influences the effect of the genetic variant or genetic score on CETP activity, then the observed effects are strictly genetic and do not represent a broadly applicable feature of CETP inhibition.

### 6 Meta-analysis of randomized controlled trials of CETP inhibitors

We conducted a sex-stratified fixed-effects meta-analysis of three RCTs of CETP inhibitors. In the REVEAL trial, rate ratios (RRs) were provided for all analyses. For ACCELERATE, the hazard ratio was provided for the main study, but RRs were reported in subgroup analyses. We calculated the RR for the main study and it was numerically identical to the hazard ratio at two decimal places. Similarly, for dal-OUTCOMES, the RR and hazard ratio were numerically identical in the main study. In sex-stratified analyses only the hazard ratios were available for this study and we could not calculate the RRs because the sex-stratified event counts were not provided. We used the hazard ratio in place of the RR for these effects. The effect estimates are summarized below along with the relevant references:

| **Study (drug)** | **Group (n; %)** | **Rate ratio (95% CI)** | **Ref.** |
|---|---|---|---|
| dal-OUTCOMES (dalcetrapib) | All (n=15,871) | 1.04 (0.93, 1.16) | [11] |
| | Male (n=12,801; 81%) | Hazard ratio: 1.07 (0.95, 1.21) | |
| | Female (n=3,070; 19%) | Hazard ratio: 0.92 (0.72, 1.16) | |
| ACCELERATE (evacetrapib) | All (n=12,092) | 1.01 (0.91, 1.11) | [12] |
| | Male (n=9,308; 77%) | 1.04 (0.93, 1.17) | |
| | Female (n=2,784; 23%) | 0.91 (0.74, 1.12) | |
| REVEAL (anacetrapib) | All (n=30,449) | 0.91 (0.85, 0.97) | [13] |
| | Male (n=25,534; 84%) | 0.90 (0.84, 0.97) | |
| | Female (n=4,915; 16%) | 0.93 (0.78, 1.11) | |

For the dal-OUTCOMES (main trial) and ACCELERATE (main trial and all subgroups), we estimated the standard error of the rate ratio on the natural log scale as $\sqrt{y_0^{-1} + y_1^{-1}}$ where $y_0$ and $y_1$ denote the number of events in the treatment arm and in the placebo arm, respectively. The 95% confidence intervals were then calculated on the natural log scale and are reported on the rate ratio scale.

The variances for the meta-analysis were calculated from the 95% confidence interval for all studies as:

$$V_i = \left( \frac{\ln(\mathrm{UCL}_i) - \ln(\mathrm{LCL}_i)}{2 \times \Phi^{-1}(0.05/2)} \right)^2$$

Where $i$ indexes studies, UCL is the upper confidence interval limit, LCL is the lower confidence interval limit and $\Phi^{-1}$ is the normal quantile function.

The meta-analysis weights were the inverse of the variances ($W_i = V_i^{-1}$). The meta analysis effect on the log scale is then calculated as:

$$\beta = \frac{\sum_i \ln(\mathrm{RR}_i) W_i}{\sum_i W_i}$$

$$Var(\beta) = \left( \sum_i W_i \right)^{-1}$$

For every group or subgroup.

Now, we denote the sex-specific meta-analysis estimates as $\beta_M$ for the male-only estimate and $\beta_F$ as the female-only estimate. We can conduct hypothesis testing of the null hypothesis of no effect using the following Wald statistics (following a 1 d.f. $\chi^2$ under the null):

$$\chi^2_C = \beta^2/Var(\beta)$$
$$\chi^2_M = \beta^2_M/Var(\beta_M)$$
$$\chi^2_F = \beta^2_F/Var(\beta_F)$$

It is also possible to test for heterogeneity in the sex-specific effects using the $\chi^2_H = \chi^2_M + \chi^2_F - \chi^2_C$ statistic having a $\chi^2$ distribution with 1 degree of freedom as described in Magi *et al.* (2010) [14]. This test is equivalent to a sex-interaction test.

### 7 Note on interaction scales

#### 7.1 Interpretation of the product interaction term in linear regression

In a linear regression model, the product interaction term represents an interaction contrast.

For example, assume the interaction model with one continous outcome $Y$ and two interacting variables $X_1$ and $X_2$.

$$\mathbb{E}(Y|X_1, X_2) = \beta_0 + \beta_1 X_1 + \beta_2 X_2 + \beta_{itx} X_1 X_2$$

We define the interaction contrast as the effect difference when both $X_1 = X_2 = 1$ compared to the sum of their individual effects ($X_1 = 1$ or $X_2 = 1$). All effects are taken using the absence of both covariables as the reference.

Concretely, the interaction contrast (IC) is:

$$\begin{aligned} IC &= Y_{11} - Y_{00} - [(Y_{10} - Y_{00}) + (Y_{01} - Y_{00})] \\ &= Y_{11} - Y_{10} - Y_{01} + Y_{00} \\ &= (\beta_0 + \beta_1 + \beta_2 + \beta_{itx}) - (\beta_0 + \beta_1) - (\beta_0 + \beta_2) + \beta_0 \\ &= \beta_{itx} \end{aligned}$$

Denoting $Y_{ij} = \mathbb{E}(Y|X_1 = i, X_2 = j)$.

Hence, the product interaction coefficient in a linear regression is the added effect attributable to the co-occurence of both risk factors in addition to the sum of their individual effects.

#### 7.2 Interpretation of the product interaction term in logistic regression

Using the same interaction of two covariables ($X_1$ and $X_2$) as in the previous section, we express the following logistic regression model on a binary outcome variable ($Y$).

$$\ln\left(\frac{p}{1-p}\right) = \beta_0 + \beta_1 X_1 + \beta_2 X_2 + \beta_{itx} X_1 X_2$$

Where $p = P(Y = 1|X_1, X_2)$.

In this model, the individual effects of $X_1$ and $X_2$ expressed as odds ratios are represented by $e^{\beta_1} = \mathrm{OR}_{10}$ and $e^{\beta_2} = \mathrm{OR}_{01}$, respectively. Their combined effect when compared to no effect of $X_1$ and $X_2$ can be derived as follows:

$$\begin{aligned}
\ln(\mathrm{OR}_{11}) &= \ln\left(\frac{\mathrm{odds}(Y|X_1 = 1, X_2 = 1)}{\mathrm{odds}(Y|X_1 = 0, X_2 = 0)}\right) \\
&= \ln(\mathrm{odds}(Y|X_1 = 1, X_2 = 1) - \ln(\mathrm{odds}(Y|X_1 = 0, X_2 = 0)) \\
&= (\beta_0 + \beta_1 + \beta_2 + \beta_{itx}) - \beta_0 \\
&= \beta_1 + \beta_2 + \beta_{itx} \\
&\rightarrow OR_{11} = e^{\beta_1+\beta_2+\beta_{itx}} = e^{\beta_1} \cdot e^{\beta_2} \cdot e^{\beta_{itx}} = \mathrm{OR}_{10} \cdot \mathrm{OR}_{01} \cdot e^{\beta_{itx}}
\end{aligned}$$

This reveals how the interaction coefficient ($\beta_{itx}$) represents a multiplicative change from the combined individual effects of the covariables on the odds ratio scale [15]:

$$e^{\beta_{itx}} = \frac{\mathrm{OR}_{11}}{\mathrm{OR}_{10}\ \mathrm{OR}_{01}}$$

#### 7.3 Estimating additive interactions from a logistic regression model

It is often argued that additive interactions are most relevant to the study of disease etiology or public health [16]. For this reason, we calculated additive interactions from the logistic regression results. We used two complementary strategies. First, we calculated the Relative Excess Risk due to Interaction (RERI) on the odds ratio scale as defined in [15]:

$$\begin{aligned}
\mathrm{RERI} &= (\mathrm{RR}_{11} - 1) - (\mathrm{RR}_{10} - 1) - (\mathrm{RR}_{01} - 1) \\
&= \mathrm{RR}_{11} - \mathrm{RR}_{10} - \mathrm{RR}_{01} + 1 \\
&\approx e^{\hat{\beta}_1+\hat{\beta}_2+\hat{\beta}_{itx}} - e^{\hat{\beta}_1} - e^{\hat{\beta}_2} + 1
\end{aligned}$$

Which holds if the odds ratio approximates the relative risk as is the case with rare outcomes. This statistic can be seen as the additive deviation on the relative risk scale due to the interaction [15]. In our analyses, the RERI were computed from the logistic regression fit using the "interactionR" R package (https://cran.r-project.org/web/packages/interactionR/index.html) and using the "mover" method to compute the confidence interval [17].

Because this measure is a deviation on the relative risk scale, we also considered interaction contrasts on the probability scale which may be more interpretable. For this analysis, we computed marginal predicted probabilities from the logistic regression fit by using a weighted average across observed covariable levels while fixing the interaction variables. We then used the interaction contrast (IC) as defined in Section 7.1:

$$IC = P_{11} - P_{10} - P_{01} + P_{00}$$

With $P_{ij} = P(Y = 1|X_1 = i, X_2 = j)$

We used the "boot" R package to construct 95% confidence intervals for this statistic using the percentile method and 2,000 bootstrap replicates.

### Supplementary Methods References


1. Bycroft, C. *et al.* The UK Biobank resource with deep phenotyping and genomic data. en. *Nature* **562,** 203–209 (Oct. 2018).
2. Legault, M. A. *et al.* A genetic model of ivabradine recapitulates results from randomized clinical trials. *PLoS One* **15,** e0236193 (2020).
3. Tardif, J. C. *et al.* Genotype-Dependent Effects of Dalcetrapib on Cholesterol Efflux and Inflammation: Concordance With Clinical Outcomes. *Circ Cardiovasc Genet* **9,** 340–348 (Aug. 2016).
4. Blauw Lisanne L. *et al.* CETP (Cholesteryl Ester Transfer Protein) Concentration. *Circulation: Genomic and Precision Medicine* **11,** e002034 (May 2018).
5. Kettunen, J. *et al.* Genome-wide study for circulating metabolites identifies 62 loci and reveals novel systemic effects of LPA. en. *Nat. Commun.* **7,** 11122 (Mar. 2016).
6. Gao, X., Starmer, J. & Martin, E. R. A multiple testing correction method for genetic association studies using correlated single nucleotide polymorphisms. en. *Genet. Epidemiol.* **32,** 361–369 (May 2008).
7. Burgess, S., Small, D. S. & Thompson, S. G. A review of instrumental variable estimators for Mendelian randomization. *Stat Methods Med Res* **26,** 2333–2355 (Oct. 2017).
8. Lousdal, M. L. An introduction to instrumental variable assumptions, validation and estimation. *Emerg Themes Epidemiol* **15,** 1 (2018).
9. Pearl, J., Glymour, M. & Jewell, N. P. *Causal Inference in Statistics: A Primer* en (John Wiley & Sons, Mar. 2016).
10. Hernán, M. A. & Robins, J. M. *Causal Inference: What If* (Chapman & Hall/CRC, Boca Raton, 2020).
